# Association of AAT/*SERPINA1* PI*Z Variant with Gestational Duration and Prevention of Premature Birth in Knockout Mice by AAT-Supplementation

**DOI:** 10.1101/2025.11.21.25340724

**Authors:** E Koivulehto, A Pasanen, AM Haapalainen, H Tiensuu, FinnGen, M Rämet, M Hallman

## Abstract

About 10% of pregnancies end prematurely before 37 weeks, without effective prevention therapies. Previously, we identified rare damaging variants in *SERPINA1* encoding Alpha-1-antitrypsin (AAT) in families with recurrent spontaneous preterm births (SPTB) and detected decreased protein and mRNA levels of AAT/*SERPINA1* from placentas in SPTB. Here, we investigated genetic associations between *SERPINA1* variants and gestational duration and evaluated AAT supplementation as a therapeutic intervention in a mouse model of preterm birth. *SERPINA1* Pi*Z variant (rs28929474-T) was associated with gestational duration (*P* < 5×10^-8^) in European-ancestry mothers with preterm and term deliveries. We detected a nine-day decrease in gestational duration and a fourfold Odds for preterm birth vs. term birth in Pi*Z homozygotes (Pi*ZZ), compared to other genotypes. In transgenic mice without endogenous AAT, exogenous Prolastina^®^ treatment inhibited LPS-induced preterm births (*P* < 0.05). Supplemented AAT was preferentially deposited in the placenta. Our findings support AAT’s protective role in SPTB and highlight its therapeutic potential, particularly in *SERPINA1* Pi*ZZ genotype carriers.

## Introduction

Annually, nearly 15 million infants are born prematurely, affecting approximately 10% of all pregnancies (Purisch & Gyamfi-Bannerman, 2017). Prematurity is associated with nearly one million neonatal deaths and potentially lifelong morbidities in survivors, such as respiratory and neurodevelopmental impairments (Chawla & Agarwal, 2022). Notably, spontaneous premature births (SPTB) account for approximately 70% of all PTB cases (Bhattacharjee & Maitra, 2021).

The etiology of SPTB is intricate and multifactorial, reflecting the heterogeneity of its underlying causes and clinical presentations (Menon, 2008, Romero et al. 2014). Emerging evidence suggests that during mid-gestation, an imbalance between pro-inflammatory and anti-inflammatory pathways may disrupt immune homeostasis, triggering SPTB (Leimert et al., 2021;Cappelletti et al., 2020; Tiensuu et al. 2019). The suggested key mediators include well-characterized cytokines and chemokines such as IL-1β, IL-6, IL-8, and TNF-α (Huang et al., 2021), as well as a myriad of anti-inflammatory proteinases, including members of the SERPIN family such as *SERPINA1* (Haapalainen et al., 2018; Tiensuu et al., 2022).

Alpha-1-antitrypsin (AAT) is a serine protease inhibitor encoded by the serine protease inhibitor (SERPIN) family member *SERPINA1* (Janciauskiene et al., 2011). AAT can neutralize neutrophil elastase (NE), which is essential for neutrophil chemotaxis and tissue remodeling (Feng et al., 2016). Under the influence of inflammatory cytokines (e.g., IL-6 and IL-8), the concentration of AAT can increase up to fourfold (O’brien et al., 2022). AAT deficiency (AATD), a rare genetic disease, has been associated with various lung-related morbidities, including asthma, emphysema, and COPD (Toumpanakis & Usmani, 2023) as well as liver diseases such as fibrosis (Mitchell & Khan, 2017). To date, over 500 *SERPINA1* variants have been reported in the ClinVar database. Among these, Pi*Z (rs28929474) and Pi*S (rs17580) are the most notorious, being associated with AATD (Annunziata et al., 2024). Case reports of patients who are either heterozygous or homozygous for these variants, i.e., carrying one or two copies of the risk allele, have demonstrated low circulating levels of AAT (<0.5 g/L) and impaired lung function (Annunziata et al., 2024; Miravitlles et al., 2023).

During pregnancy, mean serum levels of AAT gradually increase, with a marked rise by the third trimester, reaching 4–6-fold higher (3–4 g/L) than baseline. At delivery, AAT levels remain elevated compared to nonpregnant controls (Tissarinen et al. 2024). Alongside its inhibitory function in inflammation, AAT has proposed roles in endometrial angiogenesis and vascularization, as well as in trophoblast invasion and embryo implantation (Jezela-Stanek & Chorostowska-Wynimko, 2019). Low serum AAT levels (i.e., AATD) have been linked to elevated inflammatory cytokines as well as adverse pregnancy outcomes such as spontaneous abortions and PTB (Annunziata et al., 2024; Guarnieri et al., 2022; Orimoloye et al., 2024; Tiensuu et al., 2022).

Decreased expression of AAT at the basal placenta was shown to be associated with SPTB, leading to the hypothesis that reduced AAT levels may result in enhanced degradation of placental fibrinoid deposits and the release of proteins into the cervix, potentially triggering SPTB (Tiensuu et al., 2022). This finding highlights AAT augmentation as a potential treatment option for addressing imminent SPTB.

As AATD is often diagnosed after the onset of significant symptoms (Greulich & Vogelmeier, 2016), it is underdiagnosed in women of reproductive age (Greulich & Vogelmeier, 2016; Orimoloye et al., 2024). Nonetheless, AAT augmentation has been used safely to relieve lung and liver morbidities during pregnancy (Annunziata et al., 2024; Giesler et al., 1977; Guarnieri et al., 2022). Whether AATD augmentation in pregnant women with subclinical disease can reduce the risk of SPTB remains to be determined.

In the present study, we investigated the genetic and molecular biological characteristics of *SERPINA1*/AAT in the context of SPTB and explored AAT therapy as a potential intervention. We first examined whether the AAT deficiency-associated Pi*Z variant (rs28929474) is a potential gestational modulator by analyzing data from mothers who delivered preterm or at full term. To evaluate the therapeutic potential, we tested the efficacy of AAT supplementation (Prolastin^®^, Grifols) in an AATD transgenic mouse model of inflammation-induced preterm birth.

## Material and Methods

### Genetic analysis of *SERPINA1*

We tested the association of *SERPINA1* Pi*Z variant (rs28929474) with SPTB and gestational duration in mothers with term and preterm deliveries with data from the FinnGen research project DF12 (www.finngen.fi) and European-ancestry summary data contributed by the EGG Consortium (downloaded from www.egg-consortium.org; Solé-Navais et al. 2023). The association statistics for rs28929474 were obtained as part of a genome-wide association study (GWAS) meta-analysis; therefore, the alpha level of *P* <5×10^-8^ was used to denote a significant association in the original scan. The study cohorts and meta-analysis are described below.

### Phenotype definitions

In FinnGen, we defined preterm birth with the ICD codes O60 (ICD-10), 6440B (ICD-9), and ICD-8: 63497 (ICD-8), and full-term births as births that occurred at 38-42 weeks of gestation. We excluded deliveries associated with preeclampsia, polyhydramnios, and twin or multiple pregnancies. In the EGG consortium summary data, preterm birth was defined as birth before 37 completed weeks of gestation or by using the ICD-10 O60 code, and controls were defined as deliveries occurring at 39-42 weeks of gestation.

### GWAS and meta-analysis

We conducted GWAS of FinnGen data with regenie v 2.2.4 (Mbatchou et al., 2021) whole genome regression pipeline at FinnGen’s sandbox environment. Age, the top 10 principal components, FinnGen chip version (1 or 2), and legacy genotyping batch were used as covariates. Logistic regression was used for SPTB (9,822 cases and 186,977 controls), while linear regression was used for gestational duration (*N* = 67,615). Gestational duration was measured in days, and we applied a rank-based inverse normal transformation.

The EGG summary data originate from a meta-analysis of European GWA studies on SPTB (15,419 cases and 217,871 controls) and gestational duration (measured in days; *N* = 151,987). The GWAs of the participating cohorts were run with additive models, and cohort-specific QC was applied according to EGG Consortium recommendations (Solé-Navais et al., 2023). The combined effects were estimated with fixed-effects inverse-variance weighted analysis with METAL meta-analysis tool (Willer et al., 2010).

We utilized METAL (Willer et al., 2010) to conduct a meta-analysis of SPTB and gestational duration using the FinnGen and EGG data. We used fixed-effects inverse-variance weighted analysis for SPTB, and a *p*-value-based approach for gestational duration. The EGG data of SPTB includes samples from an earlier FinnGen release (R5); therefore, for the meta-analysis of SPTB, we excluded these samples from the FinnGen data. The sample number in the meta-analysis of SPTB was 344,628 (21,188 cases and 323,440 controls), and 219,602 in the analysis of gestational duration.

### FinnGen ethics statement

Study subjects in FinnGen provided informed consent for biobank research, as mandated by the Finnish Biobank Act. Alternatively, separate research cohorts, collected before the Finnish Biobank Act came into effect (September 2013) and the start of FinnGen (August 2017), were collected based on study-specific consents and later transferred to the Finnish biobanks after approval by Fimea (Finnish Medicines Agency), the National Supervisory Authority for Welfare and Health.

Recruitment protocols followed the biobank protocols approved by Fimea. The Coordinating Ethics Committee of the Hospital District of Helsinki and Uusimaa (HUS) statement number for the FinnGen study is Nr HUS/990/2017. The FinnGen study was approved by the Finnish Institute for Health and Welfare. The specific permit numbers are listed in Supplementary file 1.

### Interaction analysis

We performed gene-by-environment interaction (G × E) analysis using FinnGen data and generalized linear models (GLMs) in R, as follows: glm(gestational duration ∼ genotype * BMI + covariates, family=gaussian). The covariates were the standard FinnGen covariates, including latent principal components 1-10, age, and the genotyping chip and batch.

## Animal model studies

### Mice

C57BL/6J-Serpina1em3Chmu/J knock-out (KO) mice, lacking exon 2 from the *Serpina1a–e* genes (*Serpina1a–e* KO), were obtained from Jackson Laboratories (USA) and described by Borel et al. (2018). Mice were maintained under standard housing conditions. Primiparous *Serpina1a–e* KO and wild-type (WT) females (7–15 weeks old) were time-mated with males of the opposite genotype. Vaginal plug detection marked 0.5 days post coitum (dpc). We conducted two separate mice studies: Cohort 1 to study the efficacy of AAT supplementation and Cohort 2 to investigate the acute inflammatory response to LPS and supplementation.

### Mice Cohort 1: Efficacy of AAT in Preventing LPS-induced SPTB

At 16.5 dpc, dams were randomized by genotype and age into four treatment groups (n = 12 per group, n total = 48): 1) WT-Placebo; 2) WT-AAT; 3) KO-Placebo; 4) KO-AAT. Human alpha-1-antitrypsin (hAAT; Prolastin^®^, Grifols) was administered intraperitoneally (12.5 mg, 625 mg/kg). After 2.5 hours, lipopolysaccharide (LPS; 1.25 µg in 100 µL) was injected i.p. to induce inflammation. Control mice received saline (NaCl) in the same volume as Prolastin^®^-treated dams. Dams were monitored for labor; parturition within 48 hours post-LPS was defined as preterm.

These dams were euthanized mid-labor for sample collection. Dams not delivering by 48 hours (18.5 dpc) were euthanized, and blood was collected, and tissues were either snap-frozen or formalin-fixed.

### Mice Cohort 2: Inflammatory Response to LPS

In a parallel experiment, WT and KO dams were sacrificed at 16.5 dpc (n = 3 per group, n total = 36). Groups included: mock (untreated), AAT-or placebo-treated, and AAT-or placebo + LPS-treated. AAT/placebo-only groups were euthanized 4.5 hours post-injection. For AAT/placebo + LPS groups, LPS was given 2.5 hours after AAT/placebo, and mice were sacrificed 2 hours later (i.e., 4.5 h total post-AAT/placebo). LPS-only mice were sacrificed 2 hours post-injection.

Inflammatory response was analyzed using serum cytokine multiplex assay as described below.

### Pup assessment

Pup’s biological and developmental maturity was assessed by measuring crown–rump (CR) length. Viability of vaginally delivered pups was determined based on oxygenation (skin color, diaphragm movement), touch responsiveness, vocalization, and overall activity. Pups meeting all criteria were classified as viable. For in utero pups, viability was assessed based on placental attachment, umbilical cord blood engorgement, skin color, and response to touch through the uterine wall. All viable pups, whether live-born or in utero, were sacrificed by decapitation. Blood was collected and pooled per litter for downstream analysis. Placentas and amniotic sacs were either fixed whole in neutral formalin or snap-frozen in liquid nitrogen.

### Animal work ethical declaration

All procedures adhered to the Helsinki Declaration and EU Directive 86/609/EEC. Animal experiments were approved by the Finnish National Animal Experiment Board (ESAVI/28802/2022) and conducted following OULAC guidelines.

### hAAT concentration and half-life in mice serum

Serum samples from dams and pups were analyzed for hAAT levels (g/L) at NordLab, University Hospital Oulu (OYS), using an immunoturbidimetric assay (cut-off: 0.17–4 g/L). Each run included internal positive (human serum) and negative (untreated WT mouse serum) controls.

AAT half-life (t^1/2^) calculated using the first-order elimination model, based on the exponential decay equation:

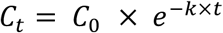

Where:

𝐶_0_ = Initial concentration at the first timepoint

𝐶_t_ = Concentration at the second timepoint

𝑡 = Time difference between the two concentrations

𝑘 = Elimination rate constant. Calculated:

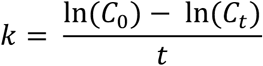

After calculating 𝑘, the half-life calculated using the following formula:

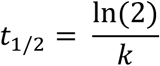

Calculations of half-life were initiated at 6 h, after serum concentrations had reached the elimination phase. Half-lives were determined for separate intervals to account for potential deviations from first-order kinetics, with an overall half-life (6–48 h) reported for reference.

### Serum cytokine multiplexing

To assess inflammatory activity, we measured 45 cytokines in serum from cohort2 dams and pups using the Mouse XL Cytokine Luminex® Performance 45-plex Fixed Panel (R&D Systems, LKTM015). Assays were performed according to the manufacturer’s instructions. Data were acquired on a Luminex MAGPIX system and analyzed with xPONENT software. The WT-mock group served as the reference.

Maternal plasma cytokine concentrations were visualized as a heatmap to compare expression across genotypes and treatments. Samples were grouped by condition (e.g., WT-MOCK, KO-LPS), and values were displayed on a diverging color scale (purple = low, white = mid, coral-red = high) to highlight shifts associated with genotype and inflammatory status.

### Histological Staining and Image Analysis

Neutral-buffered formalin-fixed tissues were embedded in paraffin and sectioned at 5 µm. Sections were deparaffinized and rehydrated before histological staining. Antigen retrieval was performed using Antigen Unmasking Solution, Tris-Based (Vector Laboratories, H-3301-250), diluted according to the manufacturer’s instructions.

Immunohistochemical (IHC) staining was carried out using the ImmPRESS^®^ HRP Horse Anti-Mouse IgG Polymer Detection Kit, Peroxidase (Vector Laboratories, MP-7402), with primary antibodies targeting either human alpha1-antitrypsin (1:1000, Sigma-Aldrich, SAB2109236) or murine alpha1-antitrypsin (1:1000, Invitrogen, PA5-79974). For each sample, negative controls omitting the primary antibody were included. Primary antibodies were incubated for 1 hour at room temperature (RT), and all further steps were conducted per the detection kit protocol.

Staining positivity and antibody localization were assessed based on the intensity of brown diaminobenzidine (DAB) coloration. Image resolution was preserved at acquisition, with brightness and sharpness enhanced post-acquisition for improved visualization.

Quantification of staining was conducted using Fiji (ImageJ) software. DAB-stained and hematoxylin-counterstained images were deconvoluted, and the DAB signal was separated for analysis. Images were converted to 8-bit format, and thresholding was applied to calculate the percentage of DAB-positive area relative to total tissue area.

### Western Blot

Western blotting was performed on placental tissues collected at 17.5 and 18.5 dpc (cohort1) to assess AAT isoforms and glycosylation status. Primary antibodies targeting human and murine AAT were used at 1:2,500 dilution. A single secondary antibody (goat anti-rabbit IgG Dylight 680 conjugate, Rockland 611-144-002-0.5) was applied to all samples at 1:10,000 dilution.

Blots were visualized using the Odyssey Infrared Imaging System (LI-COR Biosciences). GAPDH was used as a loading control. Membranes were stripped using Restore™ PLUS Western Blot Stripping Buffer (Thermo Fisher, #46430) and re-probed with anti-GAPDH antibody (1:1,000, Cell Signaling Technologies, #2118).

### RNA Extraction and RT-qPCR

Total RNA was isolated from liver and placental tissues for *Serpina1a–e* transcript analysis by RT-qPCR. Samples were obtained from time-mated KO and WT dams from both cohort1 and-2. *Eef2* was used as the reference gene.

RNA was isolated using the RNeasy Mini Kit (Qiagen, 74104) following the manufacturer’s protocol. RNA quantity and purity were assessed before cDNA synthesis, which was carried out using the Transcriptor First Strand cDNA Synthesis Kit (Roche, # 4897030001). RT-qPCR was performed on a LightCycler 96 system (Roche).

Primers and probes were obtained from Merck. Probes were labeled with FAM at the 5’ end and BHQ-1 at the 3’ end. To ensure amplification, primer and probe sequences were designed to target regions outside the knocked-out exon 2. Primer and probe sequences are as in Table 1.

**Table 1.**
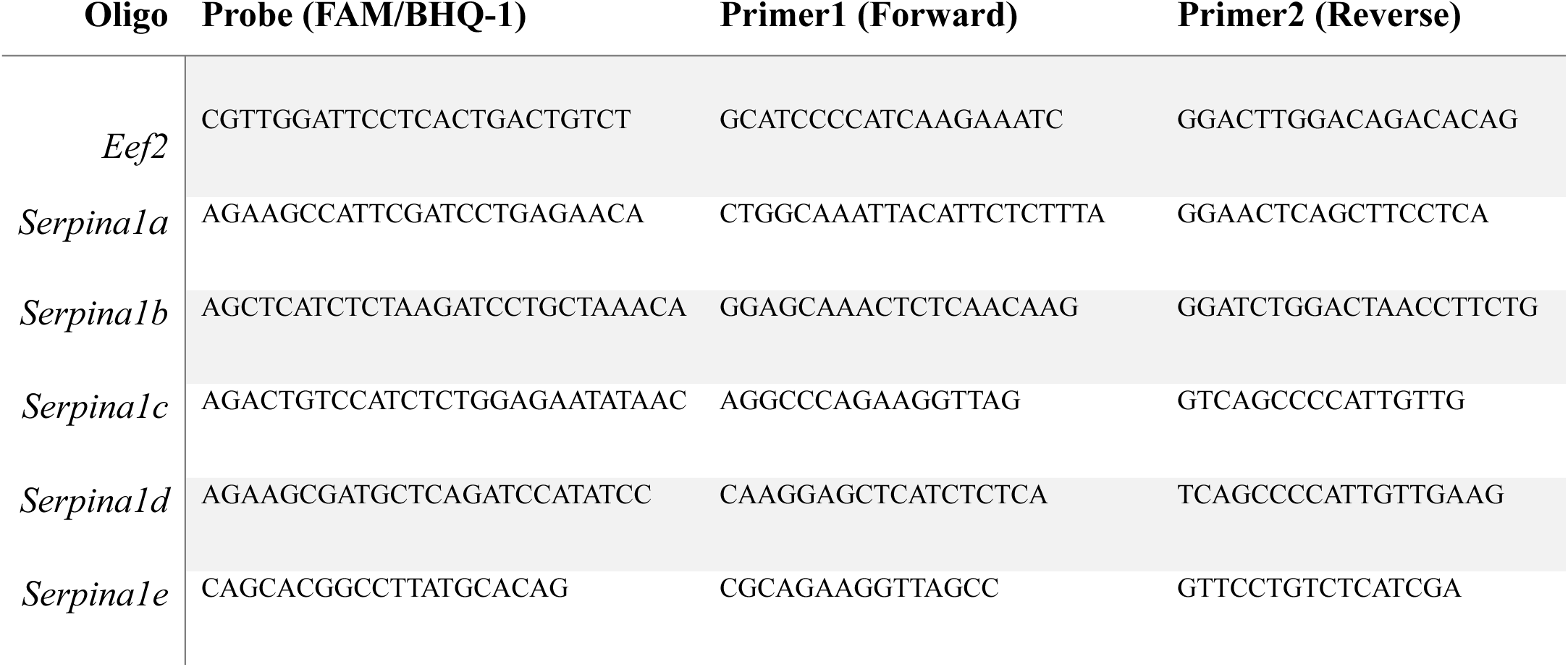
RT-qPCR primer and probe sequences.

### Mice Data Wrangling and Statistical Analysis

All data manipulation and statistical analyses were performed in R (v4.4.2; R Core Team, 2024). Data wrangling and transformation used the tidyverse (v2.0.0; Wickham et al., 2019). Plotting and alluvial diagrams of birth outcomes were generated with ggplot2 (v3.5.1; Wickham, 2016) and ggalluvial (v0.12.5; Brunson, 2020). Birth outcomes were encoded as categorical variables (“Preterm” or “Term”) and color-coded by treatment. Error bars were included for biological or technical replicates where applicable.

Normality was tested with the Shapiro–Wilk test. Due to non-normal distributions, group differences were assessed using the Kruskal–Wallis test and Dunn’s post hoc test from FSA (v0.10.0; Ogle et al., 2023), with Benjamini–Hochberg (BH) correction. Pairwise binary comparisons employed Wilcoxon rank-sum tests, also BH-adjusted. Categorical variables (e.g., gestation success, pup survival) were analyzed using Chi-square or Fisher’s exact tests, with BH-adjusted pairwise comparisons.

Survival analyses and Kaplan–Meier curves generated with survival (v3.8-3; Therneau, 2023) and survminer (v0.5.0; Kassambara & Kosinski, 2021). Global P-values were BH-adjusted.

Cytokine concentrations (pg/mL) were log₂-transformed after removing missing values and averaged across three replicates per condition. Heatmaps with hierarchical clustering were created using pheatmap (v1.0.13; Kolde, 2019), with colors ranging from dark purple (low) to dark coral (high). Significance levels were indicated as: *P* < 0.05 (*), < 0.01 (**), < 0.001 (***), and < 0.0001 (****)

## Results

### Genetic associations of *SERPINA1* with gestational duration

As rare damaging variants in *SERPINA1* have been linked to recurrent spontaneous preterm births (Tiensuu et al. 2022), we analyzed whether *SERPINA1* variants are associated with gestational duration in 219,602 European-ancestry mothers with preterm and full-term births. As shown in Figure 1, the *SERPINA1* Pi\**Z* variant (rs28929474-T) was significantly associated with decreased gestational duration (*P* = 4.33×10^-8^, Z-score =-5.477; allele frequency = 0.0327; Figure 1, Supplementary table 1). Analysis of rs28929474 genotypes in the FinnGen data revealed a nine-day difference in the median gestational duration between TT and CC genotypes (*P* = 2.53e-04 [Mann-Whitney-Wilcoxon]; Figure 2). The effect on gestational duration was more pronounced in mothers with SPTB compared to mothers with full-term deliveries (Supplementary Figure 1). This suggests that other genetic or environmental factors may modify the association between rs28929474-T and gestational duration. In the context of liver disease, high BMI was shown to amplify the association between the PI*Z allele and markers of liver damage (Hakim et al. 2021). Therefore, we conducted a gene-by-environment interaction analysis of *SERPINA1* Pi*Z variant and BMI and observed a similar pattern of worsened outcome (i.e., lower gestational duration) with increasing BMI (*P* = 0.028; Supplementary Figure 2.).

**Figure 1.**
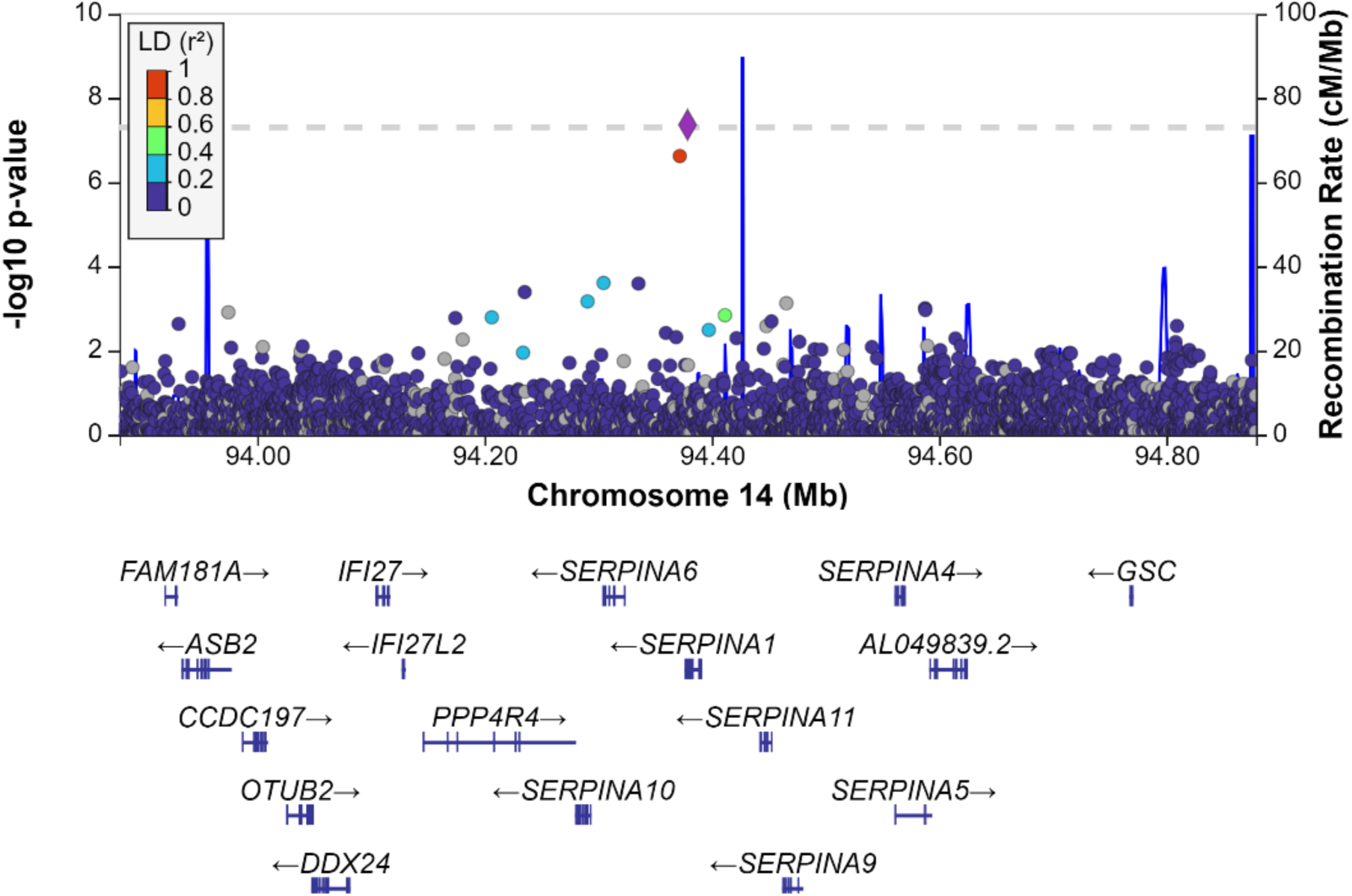
**Genetic analysis of *SERPINA1* Pi*Z variant: locus zoom-plot of rs28929474-T association with gestational duration**. Association *P*-values at the –log_10_ scale are shown on the Y-axis, and the dashed line denotes the significance threshold (*P* < 5×10^-8^; GWAS significance level used). On the X-axis, base-pair positions around the *SERPINA1* association in chromosome 14 are shown. LD (linkage disequilibrium) color indicates the strength of genetic correlation between adjacent variants.

**Figure 2.**
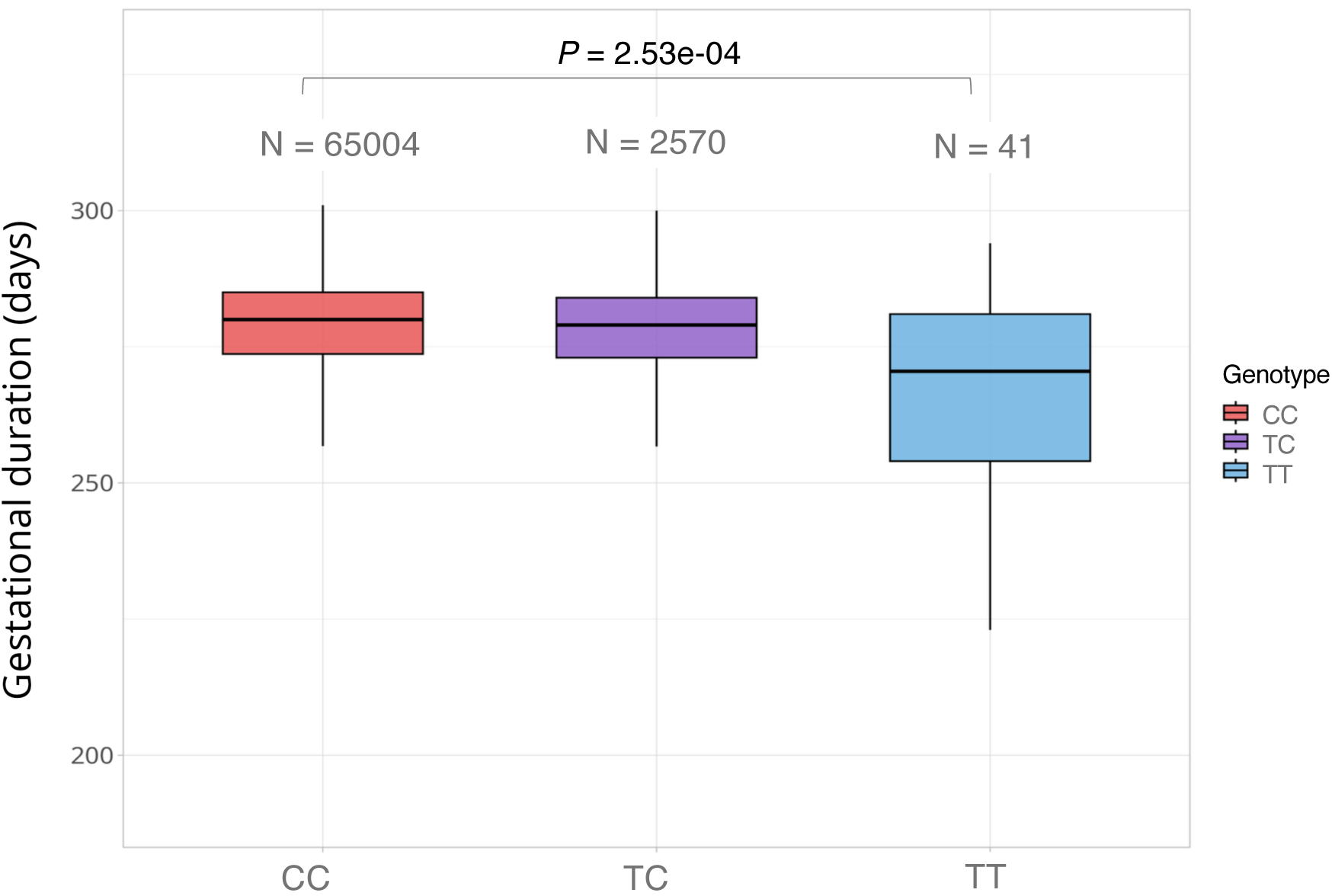
*SERPINA1* Pi*Z variant (rs28929474) genotypes and gestational duration. Considering both term and preterm births, mothers with the TT genotype (Pi*ZZ) gave birth approximately nine days earlier than mothers with the CC genotype (*P =* 2.53e-04).

The genotype association pattern of the *SERPINA1* Pi*Z variant (Figure 2) resembled that of a recessive model, i.e., people with two copies of the Pi*Z variant were significantly more affected compared to heterozygous carriers and controls. Thus, we further tested the recessive model in the association analysis of the FinnGen data and observed a larger effect size (Table 2). This indicates that the effect on shorter gestational duration and preterm birth is more potent in individuals with the *SERPINA1* PI*ZZ (rs28929474-TT) genotype. In conclusion, the *SERPINA1* Pi*Z variant appears to be a significant modifier of gestational duration in mothers of European ancestry.

**Table 2.** Association analysis of *SERPINA1* Pi*Z variant and spontaneous preterm birth. Association statistics for the additive model in the meta-analysis, EGG consortium, and FinnGen data, and association statistics for the recessive model in the FinnGen data.

| P-value | Odds Ratio (95% confidence intervals) | SPTB study |
| --- | --- | --- |
| $1.16\text{e-}04$ | 1.158 (1.083–1.232) | Meta-Analysis |
| $2.62\text{e-}02$ | 1.109 (1.018–1.202) | EGG |
| $6.16\text{e-}03$ | 1.151 (1.050–1.251) | FinnGen |
| $9.43\text{e-}06$ | 4.332 (3.684–4.981) | FinnGen, recessive model |

### AAT Supplementation prevents premature birth in knockout mice

To assess whether AAT supplementation could be used to prevent inflammation-induced preterm birth, we utilized *Serpina1a–e KO* mice and wild-type mice. In transgenic *Serpina1a–e KO* mouse strain, average gestation ranged from 19.5 to 20 days post-coitum (dpc). We used LPS to induce PTB. PTB was defined as delivery before 18.5 dpc, coinciding with the sample collection endpoint (48 hours post-LPS injection). To minimize stress, Prolastin^®^ (AAT) or placebo was administered via a single intraperitoneal (i.p.) injection. No adverse effects were observed during the 2.5-hour monitoring period prior to LPS administration.

LPS administered i.p. at 16.5 dpc triggered dose-dependent inflammatory symptoms (panting, disorientation, lethargy, eye squinting, agitation) in both WT and KO dams. A dose of 1.25 μg was selected for further studies, as it resulted in moderate inflammatory symptoms and shortened pregnancy in most, but not all, *in Serpina1a–e* KO and wild-type mice (Supplementary Figure 3).

As shown in Figure 3A, AAT efficacy in preventing inflammation-induced PTB varied by genotype (Figure 3.A–B). While AAT supplementation had no apparent effect on length of gestation upon LPS challenge in WT mice, it had 50% efficacy (6/12) in KO mice (Figure 3.A-B). Dunn’s multiple comparisons test (FDR-adjusted) showed that KO-AAT dams had significantly prolonged gestation compared to KO-Placebo (*P <* 0.032), WT-AAT (*P <* 0.047), and WT-Placebo (*P <* 0.043) (Figure 3A). AAT administered 2 hours after LPS had no protective effect; all eight (8) dams (4 WT dams; 4 KO dams) dams delivered prematurely within 20–24 hours. These data indicate that AAT supplementation protects mice with AAT deficiency from LPS-triggered PTB.

**Figure 3.**
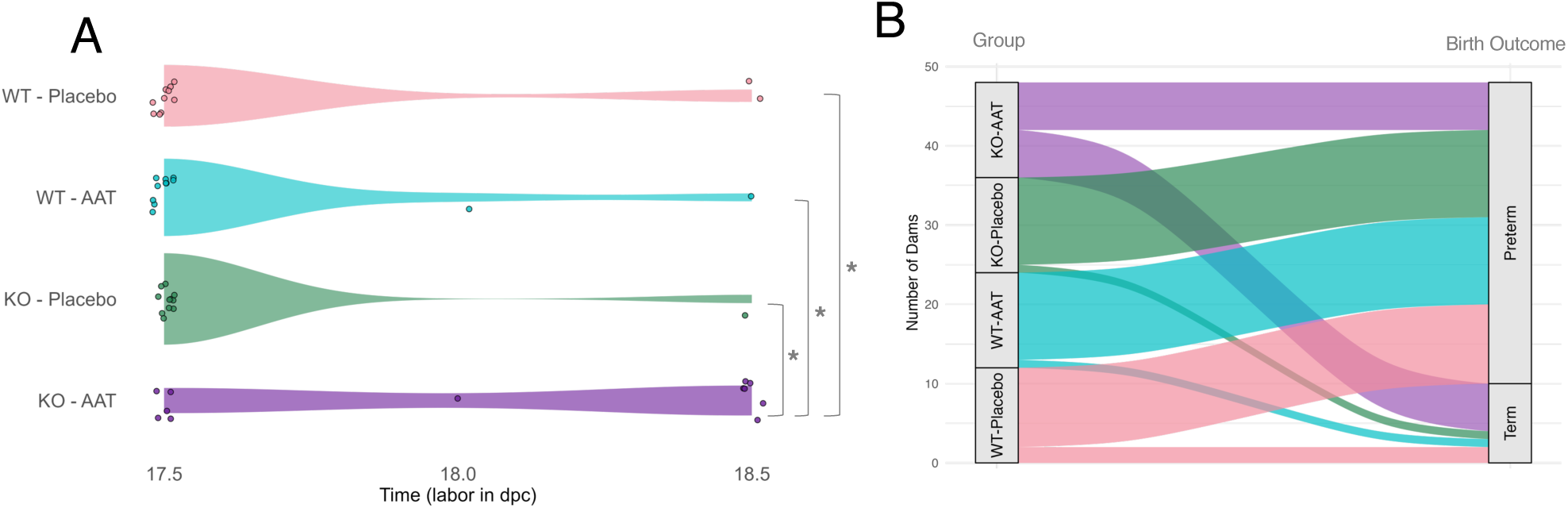
**Birth outcomes in WT and Serpina1 KO dams treated with placebo or AAT and challenged with LPS at 16.5 dpc**. (**A**) KO-AAT dams had a significant delay in labor onset compared to other groups (adjusted *Pc<* 0.05). (**B**) Alluvial plot of birth outcomes (preterm vs. term) showed no statistically significant group differences; statistical analyses were performed using Kruskal–Wallis with Dunn’s post hoc tests and Benjamini–Hochberg correction. Preterm birth PTB) was defined as delivery occurring within 48 hours after LPS challenge, whereas term birth (TB) occurred beyond this period (endpoint)

### AAT Supplementation Improves Survival Rate and Viability of Pups

Next, we evaluated the effect of AAT supplementation on fetuses. Developmental assessment using crown–rump (CR) length measurements in pups surviving beyond 48 hours—corresponding to the same dams analyzed in Figure 3 (Cohort 1) — indicated that Prolastin^®^ treatment did not impair fetal growth (Figure 4.A–B). KO-AAT litters had significantly longer CR lengths compared to both KO-placebo (*P* < 0.025) and WT-placebo litters (*P* < 0.023), indicating continued fetal growth toward term with AAT supplementation. CR lengths were consistent with expected developmental milestones and reflective of the gestational age at delivery for each litter. For pups that reached 18.5 dpc, CR growth between 16.5 and 18.5 dpc averaged approximately 0.6 cm.

**Figure 4.**
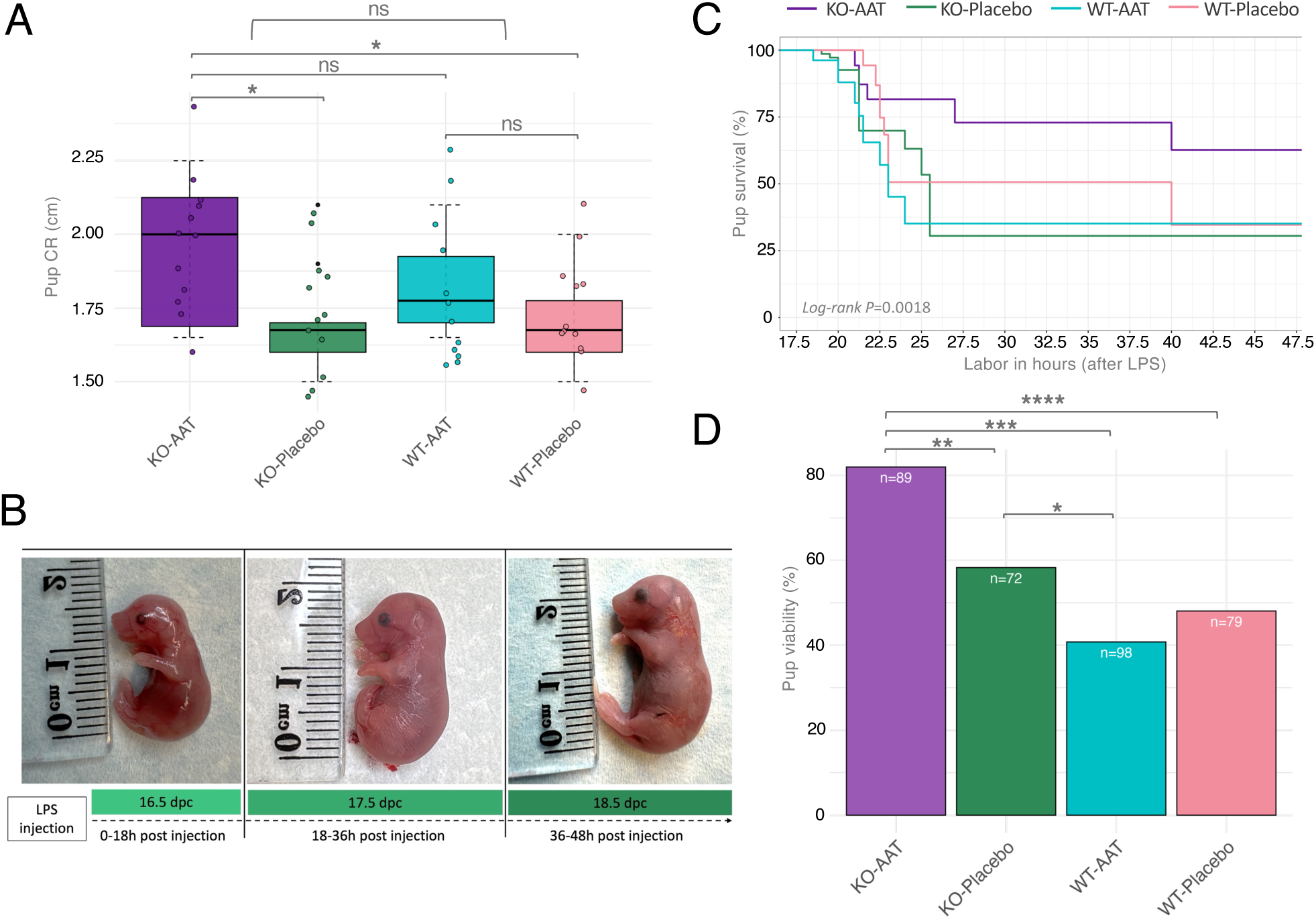
**Litter outcomes in WT and Serpina1 KO dams treated with placebo or AAT and challenged with LPS at 16.5 dpc**. **(A)** Pups from KO-AAT dams showed significantly increased crown–rump length at 18.5 dpc compared to KO-and WT-placebo groups (*, *P* < 0.025; *P =* 0.023). **(B)** Schematic presentation of developmental milestones across late gestation (from 16.5 to 18.5 dpc). **(C)** Kaplan–Meier analysis, based on binary survival data (alive = 1, dead = 0) with pups assigned to timepoints corresponding to survival duration, indicated improved overall postnatal survival in KO-AAT pups (adjusted global log-rank *P* < 0.0018). **(D)** The same binary survival data were summarized by genotype–treatment group to calculate overall postnatal viability, presented as bar plots (**** *P* < 9.4e–08; *** *P* < 2.3e–05; ** *P* < 0.003), with KO-Placebo pups also outperforming WT-AAT (*, *P* < 0.045).

KO pups treated with Prolastin^®^ showed the highest overall survival probability (Figures 4.C–D). Global survival analysis revealed significant differences among groups (adjusted log-rank *P <* 0.0018). Following euthanasia of the dam after the delivery of the first pup, most remaining in utero pups were viable (Supplementary Figure 4.A), showing intact placental attachment and umbilical vein engorgement. In contrast, pups delivered vaginally before 18.5 dpc were largely nonviable, regardless of treatment (Supplementary Figure 4.B). Consistent with the survival analysis, litters from KO dams treated with Prolastin^®^ exhibited the highest viability percentages, with all phenotype comparisons reaching statistical significance (*P<* 9.4e–08; *P <* 2.3e–05; *P <* 0.003; Figure 4.D).

These findings suggest that selected dose of AAT is not toxic and does not compromise fetal growth. Instead, mortality in preterm-born pups likely reflects insufficient developmental immaturity at the time of delivery.

### Clearance of Human Alpha-1 Antitrypsin in Mice is Influenced by Inflammation and *Serpina1a-e* genotype

Next, we studied the kinetics of hAAT clearance after supplementation. A single 12.5mg i.p. dose of Prolastin^®^ elevated circulating hAAT to levels approximating the murine physiological range observed in non-gravid, unstimulated mice (∼2 g/L; Figure 5A). The end-point serum half-life averaged ∼38 hours post-injection. Dose of AAT was calculated based on the basal weight of young adult female mice (∼20 g) and adjusted for reduced systemic absorption associated with the *i.p*. route. Maternal weight gain during pregnancy was not factored in dose determination.

**Figure 5.**
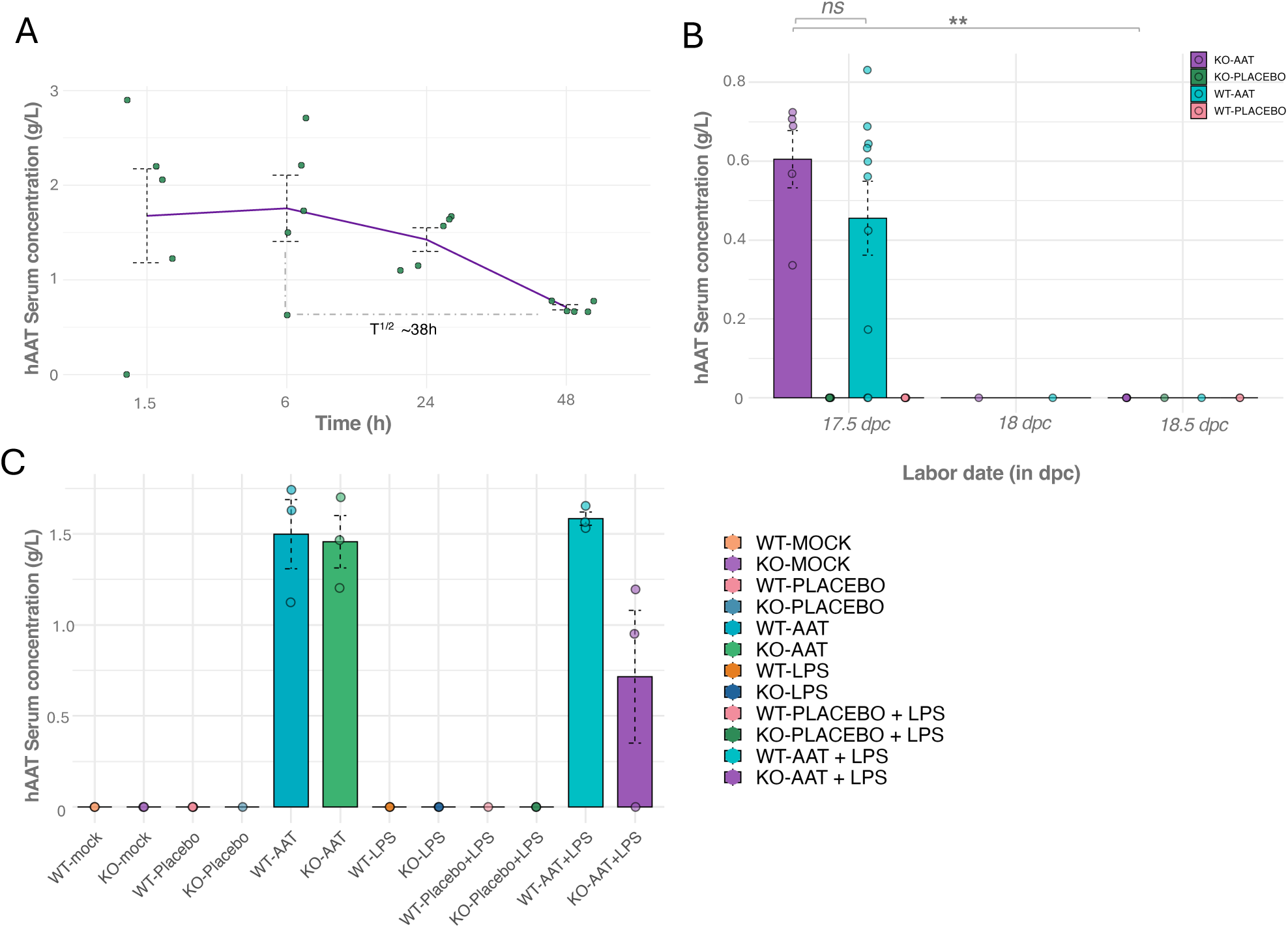
Serum levels and pharmacokinetics of human alpha-1 antitrypsin (hAAT) in Prolastin®-treated *Serpina1a-e* KO and WT mice. (**A**) In non-pregnant KO mice, hAAT peaked by 6 h and declined with a terminal half-life of ∼38 h; (**B**) in pregnant dams treated with Prolastin^®^ and challenged with LPS at 16.5 dpc, KO-AAT mice showed a significant drop in hAAT at 17.5 vs. 18.5 dpc (P *=* 0.0014), with no significant difference between WT-AAT and KO-AAT; (**C**) at 16.5 dpc, LPS reduced hAAT levels in KO-AAT (1.49 g/L vs. 1.07 g/L, ns) but not in WT-AAT dams, likely due to excess endogenous and exogenous AAT.

Prolastin^®^-treated gravid dams (Cohort 1), exposed to LPS, exhibited accelerated hAAT clearance compared to non-gravid, unstimulated mice from a separate control group (Figure 5.B). 24 hours after administration of AAT at 16.5 dpc, mean serum hAAT concentrations were similar between KO-AAT (0.50 g/L, median: 0.63 g/L) and WT-AAT dams (0.51 g/L, median: 0.58 g/L). hAAT levels became undetectable by 48 hours (18 dpc) in both groups (KO-AAT, *P <* 0.0014; WT-AAT, not significant), indicating rapid systemic clearance.

To further assess the kinetics of hAAT clearance, serum hAAT levels were measured 4.5 hours after Prolastin^®^ administration (Cohort 2) with or without LPS (Figure 5.C). Mean hAAT levels were 1.46 g/L in WT dams and 1.49 g/L in KO dams, and 1.58 g/L (WT) versus 1.07 g/L (KO) when co-administered with LPS. These data suggest that LPS challenge reduces circulating hAAT in KO mice; however, formal statistical testing was not performed due to low sample size of 3 per timepoint. Notably, WT dams maintained relatively high hAAT levels despite poor outcomes, suggesting that elevated circulating hAAT alone is insufficient to prevent inflammation-induced SPTB.

Serum AAT levels were undetectable in pups born to Prolastin^®^-treated WT and KO dams, indicating that supplementary hAAT does not readily cross the placenta to enter the fetal circulation at physiologically relevant levels (Supplementary Figure 5).

### LPS Administration results in acute systemic inflammation regardless of AAT supplementation

To evaluate the immunomodulatory effects of Prolastin^®^ (AAT) supplementation, we measured a broad panel of cytokines and growth-related mediators in Cohort 2 dams, two hours post-LPS stimulation (or in unstimulated controls) at 16.5 dpc. LPS exposure increased pro-inflammatory, anti-inflammatory, and growth-associated cytokines in both WT and KO dams (Figure 6.A–C: 7–12; Supplementary Figure 6-7). A single outlier in the KO-LPS group displayed low cytokine levels, possible due to incomplete LPS exposure (Figure 6.A–C: 8).

**Figure 6.**
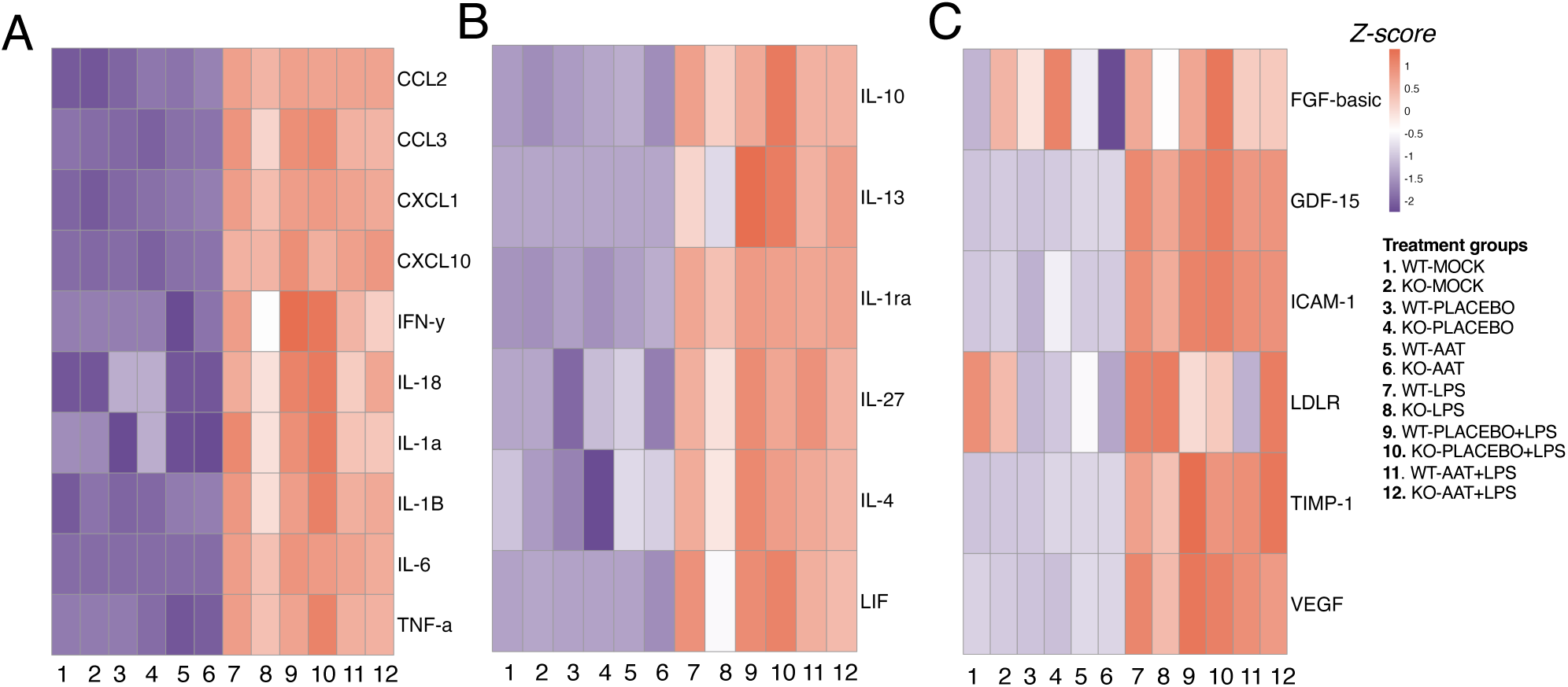
Heatmap visualization of cytokine and growth factor responses following placebo/AAT with or w/o LPS exposure at 16.5 dpc. Heatmaps showing z-score–normalized maternal serum levels of (A) pro-inflammatory, (B) anti-inflammatory, and (C) growth-associated cytokines and chemokines at 16.5 dpc in WT and *Serpina1a-e* KO dams treated with placebo or AAT, with or without LPS exposure (sample number 1-12); LPS robustly induced all cytokine classes, with no significant suppression by AAT. Sample group 8 (KO+LPS) represents a low-cytokine outlier, likely due to incomplete LPS exposure.

Among pro-inflammatory cytokines, IL-6, TNF-α, IL-1α, IL-1β, and IFN-γ were significantly elevated, along with chemokines CXCL1 and CCL2 (MCP-1), suggesting neutrophil and monocyte recruitment. Increases in IL-18, CXCL10, and CCL3 (MIP-1α) further indicated leukocyte activation, consistent with TLR4 activation via MyD88 and TRIF pathways. AAT supplementation did not significantly reduce these responses relative to placebo.

LPS also upregulated anti-inflammatory cytokines, including IL-10, IL-1ra, IL-4, IL-13, LIF, and IL-27 (Figure 6.B: 7–12), likely reflecting a compensatory mechanism to limit inflammation. While variability was observed across LPS-treated animals, AAT treatment did not affect the expression of these markers beyond levels seen in placebo-treated dams.

In addition, angiogenic and tissue integrity markers—including VEGF, ICAM-1, GDF-15, FGF-basic, TIMP-1, LDLR, and EGF—were elevated following LPS exposure (Figure 6.C: 7–12), with ICAM-1, VEGF, TIMP-1, and GDF-15 showing the most consistent upregulation. AAT had no significant effect on the expression of these growth-associated factors.

A broader cytokine and chemokine panel (Supplementary Figure 8.A) confirmed general immunomodulatory trends in dams. LPS exposure significantly increased colony-stimulating factors (M-CSF, G-CSF, GM-CSF), T cell–associated cytokines (IL-17A, IL-21), and other inflammatory mediators. Prolastin^®^ did not have a significant modulatory effect on these values.

Overall, LPS triggered a systemic inflammatory response involving both immune and tissue remodeling pathways. AAT supplementation did not significantly alter individual cytokine levels. Hence, AAT’s protective effects against LPS-induced PTB in KO dams, and its lack of efficacy in WT dams, suggest alternative mechanisms, such as placental stabilization, in the prevention of inflammation-driven preterm birth.

Fetal (pup) serum cytokine profiles were also analyzed across all groups (Supplementary Figure 7.B). Unlike the maternal compartment, fetal cytokine levels remained largely unchanged by genotype or treatment. Key inflammatory markers (e.g., IL-6, IL-1β, IL-10, CXCL10, VEGF) remained at baseline, indicating that acute maternal inflammation did not lead to a systemic fetal immune response. Of note, WT-mock-treated pups had the highest cytokine levels across analytes (Supplementary Figure 8.B:1), likely reflecting developmental maturation rather than inflammatory activation. This suggests that maternal inflammation is compartmentalized, with the placenta effectively shielding the fetus from systemic immune activation. These findings support the hypothesis that improved pregnancy outcomes in KO-AAT dams—and the lack of benefit in WT-AAT dams—are driven by maternal or placental mechanisms rather than fetal immune modulation.

### Genotype-Specific Modulation of Endogenous AAT in the Placenta and Fetal Membranes

To assess the localization and degradation of endogenous mouse AAT (mAAT) and exogenous human AAT (hAAT, Prolastin^®^), we performed immunohistochemistry (IHC) and Western blot (WB) on Cohort 1 placentas collected at 17.5 and 18.5 dpc (Figure 7.A–C). The basal plate (BP) included decidua and junctional zone, while the chorionic plate (CP) corresponded to the labyrinth layer. AAT distribution was also evaluated in the amniotic membranes (amnion and the yolk sac; here referred as AMS).

**Figure 7.**
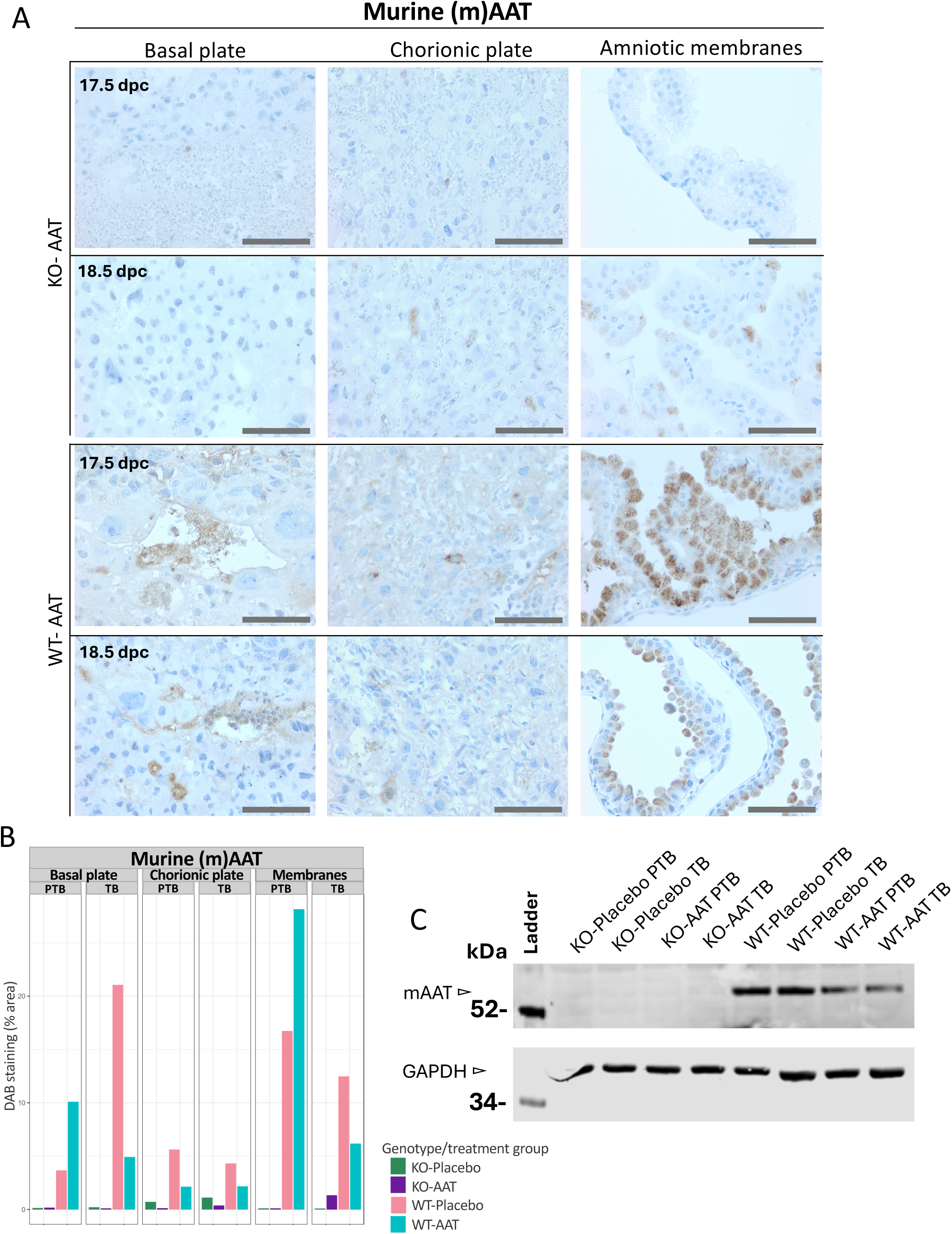
Placental immunohistochemistry and Western blot analysis of endogenous murine AAT (mAAT) expression in WT and *Serpina1a-e* KO dams treated with placebo or Prolastin^®^, and LPS. (A) IHC images (40×) show mAAT localization in the basal plate, chorionic plate, and amniotic sac at 17.5 (preterm birth; PTB) and 18.5 dpc (term birth; TB). (**B**) mAAT-positive area (% DAB staining) was quantified by region, treatment, and outcome. (**C**) Western blots of mAAT and GAPDH show expression patterns across genotypes and birth outcomes. Prolastin^®^ treatment decreased mAAT in WT treated dams at term.

Endogenous mAAT, detected by anti-mAAT IHC, showed genotype-and treatment-dependent patterns (Figure 7.A–B). In KO dams, mAAT localized mainly to the CP and AMS, being derived likely from heterozygous fetal tissues (Figure 7.A-B; Supplementary Figure 9).

Placebo-treated WT dams exhibited strong mAAT staining across CP, BP and AMS (Supplementary Figure 9; Figure 7.B), localized in blood vessels, extracellular matrix, and intracellular compartments. In Prolastin^®^-treated WT dams, mAAT staining was elevated at 17.5 dpc but decreased by 18.5 dpc, reflecting dynamic regulation in response to supplementation (Figure 7.A-B). AMS staining was higher in WT dams, with possible mAAT aggregation at 17.5 dpc, reflecting either a fetal or maternal response to LPS.

WB analysis of placental lysates (Figure 7.C) confirmed these findings: WT dams showed a consistent mAAT (∼54 kDa, fully glycosylated isoform) across treatments and timepoints. No partial or unglycosylated forms were detected, implying limited de novo synthesis or rapid glycosylation under inflammation. KO placentas lacked mAAT, supporting minimal fetal-to-maternal transfer. WT liver samples confirmed systemic AAT upregulation, while KO livers showed no detectable mAAT (Supplementary Figure 10.A).

Overall, endogenous AAT production and retention appear highly dynamic, influenced by genotype and exogenous AAT exposure. The observed reduction in IHC staining and altered isoform abundance suggest that exogenous hAAT may transiently suppress endogenous mAAT expression and affects its post-translational modifications. This may, in part, explain why AAT fails to inhibit LPS-induced preterm birth in WT mice.

### Endogenous AAT Is Retained at the Maternal-Fetal Interface Following hAAT Supplementation

To assess the localization and quantity of exogenous human AAT (hAAT, Prolastin^®^), we performed anti-hAAT IHC and WB analyses corresponding to those used for endogenous murine AAT above (Figure 8.A–C). As expected, no hAAT staining was observed in placebo-treated WT or KO dams (Supplementary Figure 9; Figure 8.B). Prolastin^®^-treated groups demonstrated genotype-dependent patterns of hAAT localization at 17.5 and 18.5 dpc (Figure 8.A–B). In KO tissues, hAAT localized to vascular and extracellular regions of both the basal plate (BP) and chorionic plate (CP), remaining detectable through 18.5 dpc. Conversely, WT placentas exhibited lower hAAT signals, consistent with reduced retention or faster clearance, possibly due to functional hepatic pathways or competitive inhibition by endogenous mAAT.

**Figure 8.**
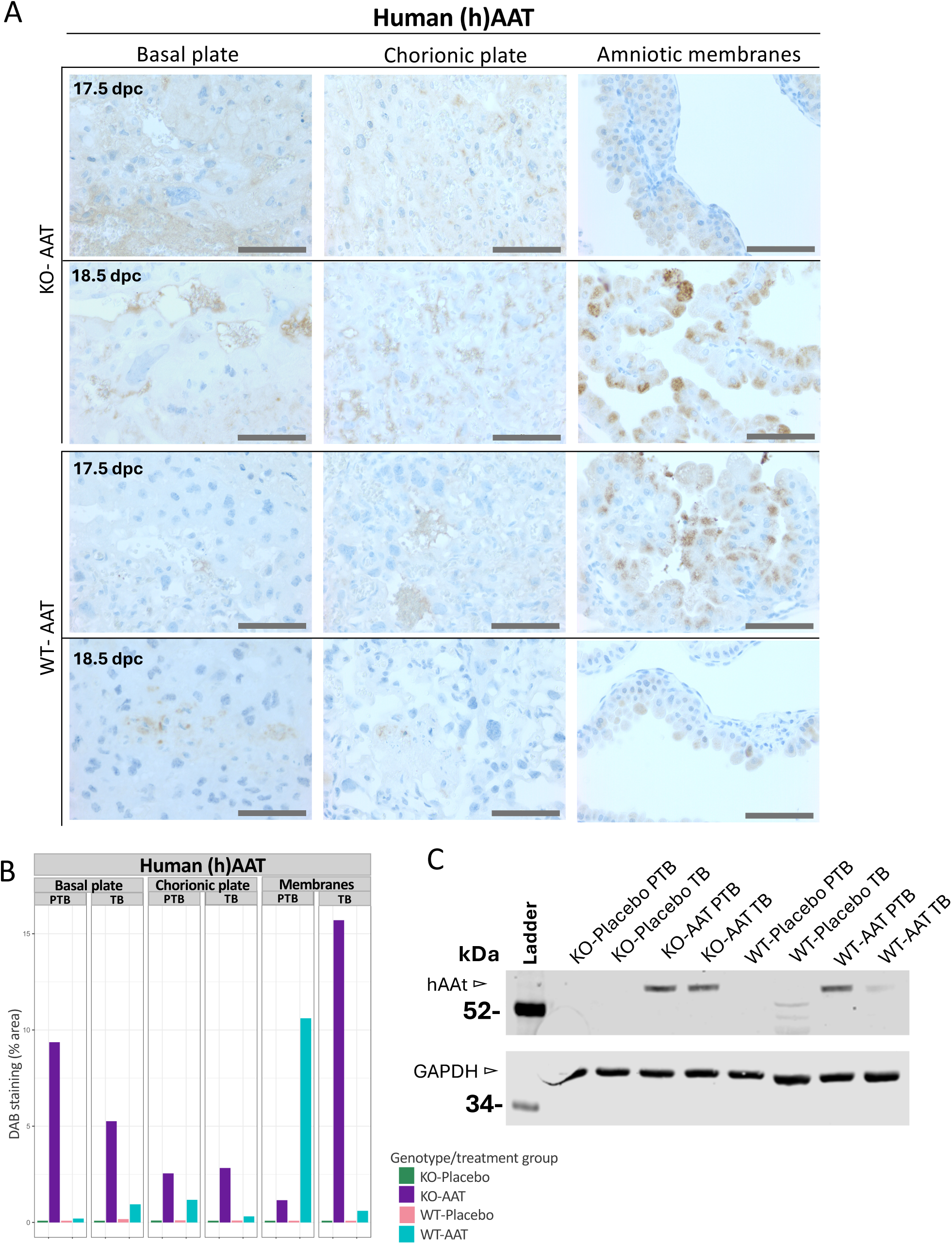
Placental immunohistochemistry and Western blot analysis of exogenous human AAT (hAAT) in WT and *Serpina1*a-e KO dams treated with placebo or Prolastin^®^, and LPS. (**A**) IHC (40×) showing hAAT localization in basal plate, chorionic plate, and amniotic sac at 17.5 and 18.5 dpc. (**B**) quantification of hAAT-positive area (% DAB staining) by region, treatment, and birth outcome. (**C**) Western blots of hAAT and GAPDH (loading control from Figure 7.C) across genotypes and birth outcomes. In combination, results show faster clearance of hAAT in WT dams and prolonged retention in KO dams, with minimal detection of hAAT in WT placental lysates at term.

hAAT staining persisted longer in KO-AAT/Prolastin^®^ dams, with visible signal at 18.5 dpc, compared to WT dams. Detected hAAT in the amniotic membranes (AMS) likely originates from maternal uterine lining and circulation rather than pup circulation (Figure 8.A-B), differing from the fetal source of mAAT (Figure 7.A-B). This suggests that exogenous hAAT accumulates more efficiently in the absence of endogenous mAAT indicating tight control of AAT levels in placental tissues.

Western blot analysis confirmed placental retention of hAAT but showed no detectable uptake in the liver (Supplementary Figure 10.B). This is consistent with localized placental accumulation. hAAT appeared only at molecular weight of approximately 54 kDa, indicating fully glycosylated, mature protein. These results validate effective delivery of hAAT to the maternal-fetal interface with limited systemic distribution following intraperitoneal administration.

In summary, combined IHC and Western blot analyses demonstrate that endogenous AAT expression is dynamically modulated by exogenous Prolastin^®^, with notable genotype-dependent differences in localization, retention, and glycosylation status. Exogenous hAAT accumulates at the placenta, highlighting its targeted bioavailability. The differencing retention patterns between KO and WT dams emphasize the critical role of endogenous AAT in clearance and tissue distribution of therapeutic AAT.

### *Serpina1a–e* mRNA Levels Are Modulated by Genotype, Inflammation, and Prolastin^®^ Supplementation

To further evaluate the effects of exogenous hAAT on AAT regulation, the expression of *Serpina1a-e* transcripts was assessed by RT-qPCR to determine the impact of exogenous AAT on *Serpina1a-e mRNA expression* during acute inflammation in liver samples from Cohort2. LPS-treated WT dams (placebo or Prolastin^®^) showed broad downregulation of *Serpina1a–e* mRNA compared to mock.

The lowest transcript levels were in WT-AAT/Prolastin^®^+LPS animals, indicating tight regulation of AAT levels during acute inflammation.

In mock-treated mice, WT livers expressed all *Serpina1* paralogs, with *Serpina1a* being the highest, while KO livers showed virtually no expression, consistent with the deletion (Figure 9.A–E).

**Figure 9.**
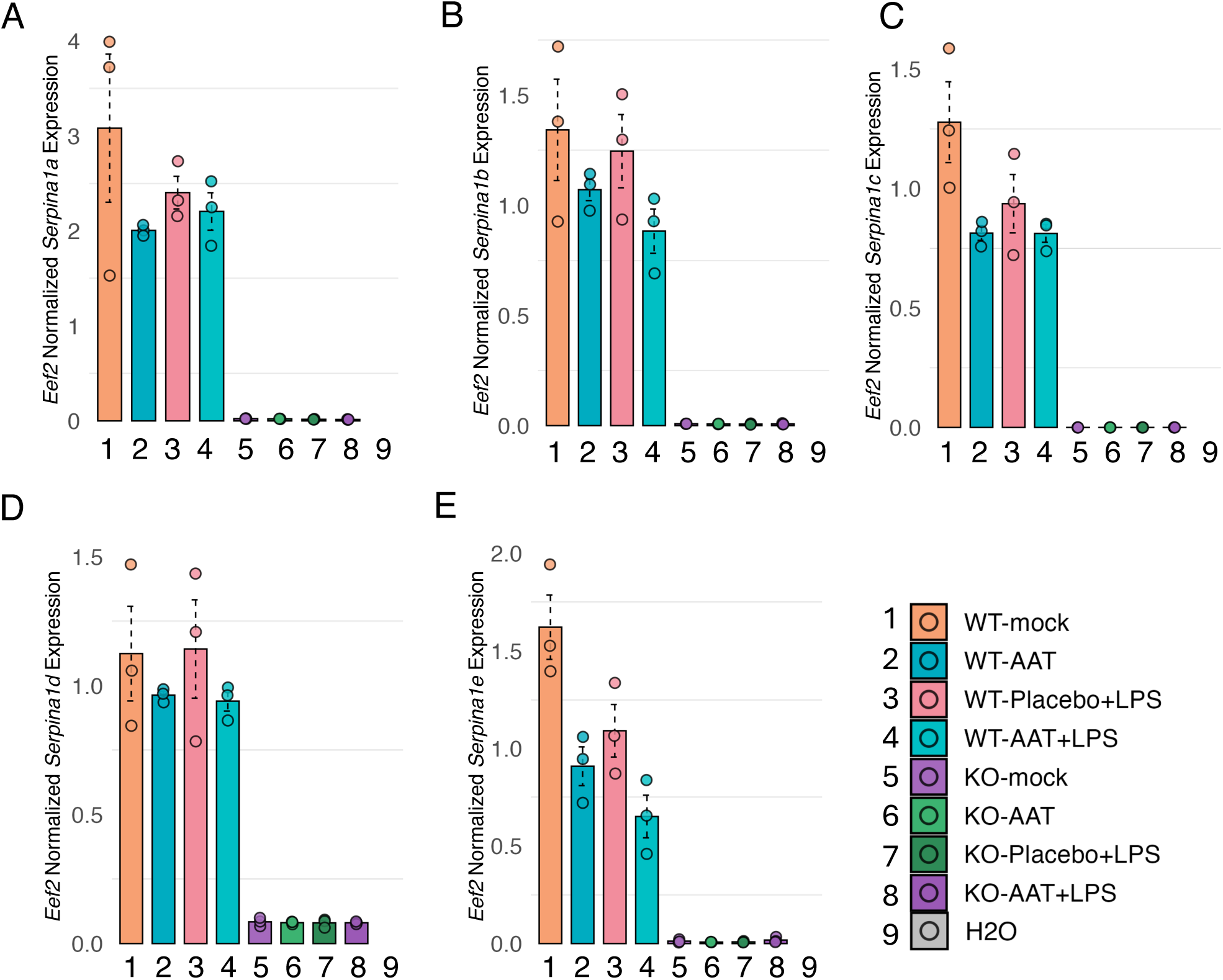
**Relative expression of *Serpina1a–e* transcripts in liver from WT and *Serpina1a–e* KO dams at 16.5 dpc (Cohort2)**. Plots (**A–E**) demonstrate individual *Serpina1* paralogs *a–e* expression by treatment group (1-8) normalized to *Eef2*. WT groups exhibited robust expression of all five paralogs downregulated by Prolastin^®^ treatment and/or LPS, with lowest levels in combined treatment (group 4). KO groups showed minimal or absent transcripts, confirming knockout, with negligible *Serpina1d* signal in KO samples.

Following Prolastin^®^ treatment, WT livers displayed reduced *Serpina1a–e* expression, indicating suppression of endogenous transcription by exogenous AAT. KO livers had minor *Serpina1d* mRNA expression, consistent with the absence of mAAT protein (Supplementary Fig. 10.A; Fig. 7.C).

### Tissue-Specific *Serpina1a-e* Transcript Profiles at Preterm and Term

To study relative roles of placenta and liver for AAT production, we assessed *Serpina1a–e* transcript levels in liver and placental samples at 17.5 dpc (PTB) and 18.5 dpc (TB) from Cohort1 (Figure 10.A–E). In WT livers, *Serpina1a–e* transcripts were downregulated in PTB dams treated with placebo (Figure 10.A–E:1), consistent with earlier data (Figure 9.A–E:3). By TB, *Serpina1a–e* expression was restored (Figure 10.A–E:2). In contrast, WT dams treated with AAT/Prolastin^®^ showed elevated transcripts at PTB (Figure 10.A–E:3) but decreased levels by TB (Figure 10.A–E:4), indicating an initial transcriptional boost followed by inhibition by exogenous AAT. KO dams showed no or minimal expression of *Serpina1a–e* (<0.13 normalized ratio; Figure 10.A–E:5–8), consistent with exon 2 deletions across all paralogs.

**Figure 10.**
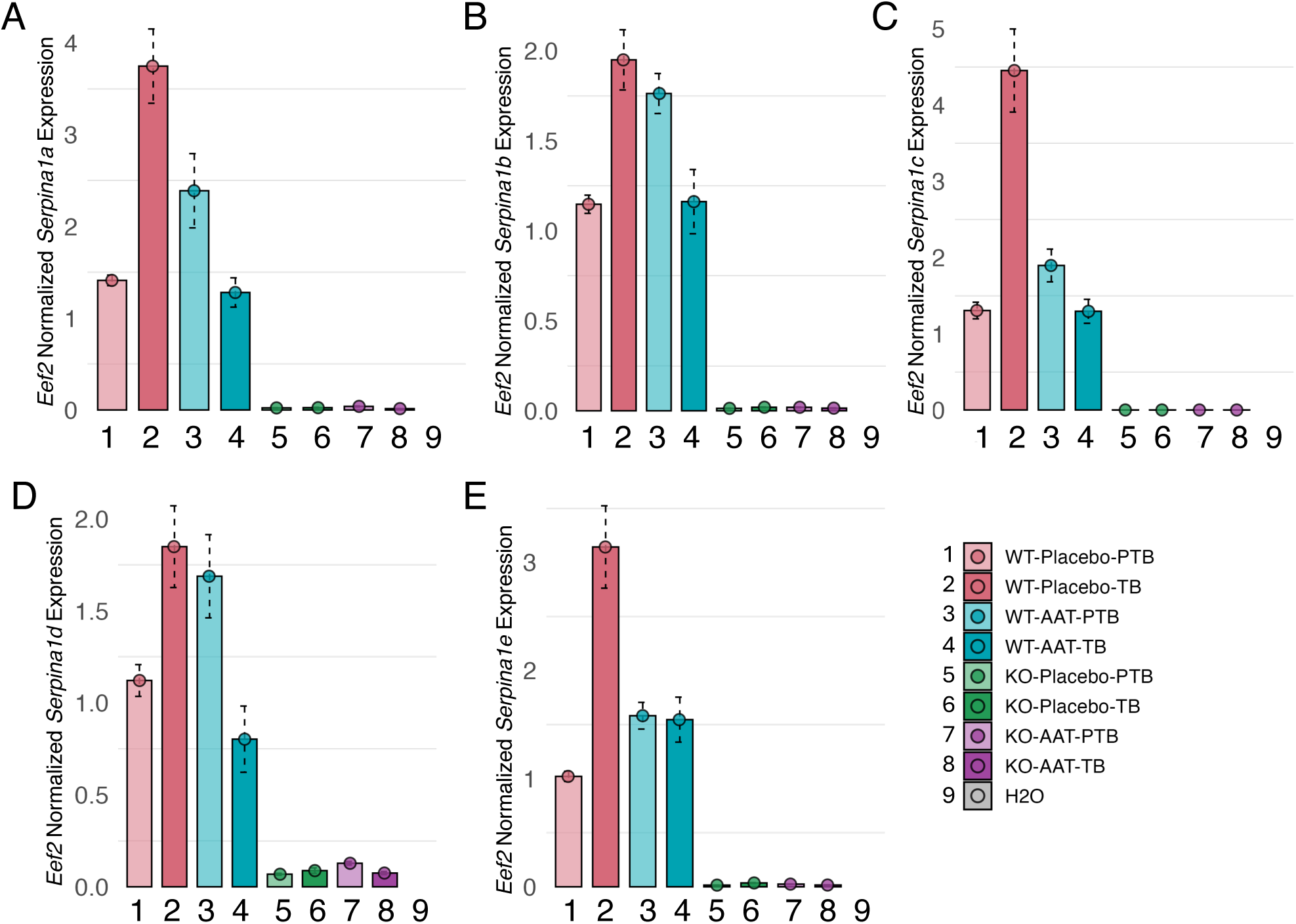
Liver mRNA expression of *Serpina1a–e* paralogs at 17.5 (PTB) and 18.5 dpc (TB) in LPS-challenged WT and KO dams (Cohort1). Plots (A–E) demonstrate individual *Serpina1* paralogs *a–e* expression by treatment group (1-8) normalized to *Eef2*. Prolastin^®^ treated WT dams at 18.5dpc (group 4), showed downregulated transcripts, while WT-Placebo groups (1-2) showed upregulated expression pattern after downregulation, suggesting feedback regulation, and diminishment of inflammatory stimuli. KO dams lacked detectable functional transcripts. Error bars demonstrated standard error between triplicates.

In placentas, *Serpina1a–e* transcripts were undetectable across groups (Supplementary Figure 11), confirming negligible local expression. These findings indicate that murine AAT is liver-derived and systemically delivered to the placenta, highlighting placental reliance on hepatic AAT for homeostasis. Consequently, systemic AAT deficiency may increase the vulnerability of placental structures.

## Discussion

Our study provides evidence linking the *SERPINA1* Pi*Z variant (rs28929474) to gestational duration and the risk of preterm birth (PTB) in humans and demonstrates that alpha-1-antitrypsin (AAT) supplementation inhibits LPS-induced spontaneous preterm birth (SPTB) in *Serpina1a-e* knockout (KO) mice. These findings emphasize AAT’s protective role in inflammation-driven pregnancy complications and highlight its therapeutic potential.

Our genetic meta-analysis of the *SERPINA1* PI*Z variant, using data from the FinnGen research project and the EGG consortium, identified a significant association between rs28929474 and gestational age. Follow-up analysis in the FinnGen data revealed a large genotypic effect between the Pi*ZZ genotype and SPTB. Mothers who were homozygous for the Pi*Z allele exhibited significantly shorter gestational duration compared to heterozygous or non-carrier individuals.

Among the women who gave birth preterm, the reduction in gestational duration was approximately 10 days in for those with Pi*ZZ genotype, compared to women with other genotypes (242 vs. 252 days). The effect of the Pi*ZZ genotype became more pronounced with increasing BMI. Interestingly, a similar pattern of worsened outcome was previously detected in gene-environment interaction analysis in the context of Pi*ZZ variant and liver disease. The substantial effect of *SERPINA1* Pi*Z alleles on gestational duration could arise from a genetic susceptibility to inflammation-induced PTB mediated by AAT deficiency. This is in consort with our previous findings of the role of AAT/*SERPINA1*at the basal placenta (Tiensuu et al., 2022).

Our mouse model analyses revealed genotype-and treatment-dependent differences in AAT processing and distribution. Exogenous human AAT given intraperitoneally localized in the placenta, rather than liver, suggesting localized action at the maternal–fetal interface. WT dams treated with Prolastin^®^ showed increased clearance of human AAT compared to KO dams, indicating an endogenous feedback loop. Both AAT deficiency and excess may be detrimental, supporting a threshold effect, further evidenced by downregulation of all *Serpina1a–e* paralogs after AAT administration and LPS challenge (Cosio et al., 2016).

Although AAT has been shown to inhibit NF-κB signalling (Yuan et al., 2022), cytokine profiling revealed no detectable suppression of acute inflammatory markers within 2 hours post-LPS. Placebo-and Prolastin^®^-treated mice exhibited similar proinflammatory cytokine profiles, suggesting AAT’s protective effects may involve longer-term modulation of immune responses, maintenance of placental integrity, and vascular remodelling (Janciauskiene et al., 2011; Pini et al., 2022; Tiensuu et al., 2022). These findings align with the multifactorial nature of SPTB (Bhattacharjee & Maitra, 2021; Goldenberg et al., 2008; Hallman et al., 2019)

In placebo-treated WT-dams, LPS induced rapid downregulation of hepatic *Serpina1a–e* transcripts (Figures 9–10, 16.5 and 17.5 dpc), followed by restored expression by 18.5 dpc, likely a feedback mechanism restoring homeostasis. In contrast, the transcript suppression continued by 18.5 dpc in Prolastin^®^-treated animals, suggesting augmentation of endogenous feedback inhibition by exogenous AAT. This points to tightly regulated AAT homeostasis during pregnancy, modulated by endogenous and therapeutic factors. Future studies should utilize spatial transcriptomics or single-cell RNA-seq to provide detailed placental cell responses and employ advanced kinetic modelling or labelled AAT to distinguish exogenous vs. endogenous sources.

LPS challenge elevated cytokine expression and induced labor; AAT supplementation improved pregnancy outcomes in KO dams but did not affect expression of inflammation markers. This implies that other factors such as maternal immune programming and maintenance of structural integrity of placenta may influence the outcome (Preston et al., 2024; Wei et al., 2024). AAT’s anti-protease effects may represent one facet of broader protective mechanisms.

Though our focus was on preventing LPS-induced SPTB, AAT supplementation may also benefit fetal neurodevelopment. Prior work links intrauterine inflammation to neurodevelopmental impairments (O’Loughlin et al., 2017). While pups were heterozygous, inflammation-induced growth restriction and neurodevelopmental risks persist. The potential of AAT to protect fetal growth and neurodevelopment, possibly by mitigating fetal brain proinflammatory signalling or preserving placental barrier integrity, warrants further investigation. Zhang et al. (2023) demonstrated, that AAT crosses the blood–brain barrier (BBB) in pups and alleviates hypoxia–ischemia brain injury, reducing BBB permeability, caspase-3 activation, neuronal death, and microglial activation, supporting its neuroprotective promise during gestation.

We acknowledge the limitations of this study. These include the obvious interspecies differences in utero-placental physiology, anatomy, regulation of the length of pregnancy and the biology of AAT. Murine endogenous AAT levels are higher and less gestationally regulated than humans (Annunziata et al., 2024; Lamontagne et al., 1981). To achieve therapeutic levels, we used a single high dose (>600 mg/kg), exceeding human augmentation therapy doses (60 mg/kg) (Campos et al., 2019). The dose achieved near-physiological murine serum levels (∼2 g/L; see Figure 4A-B) (Ostermann et al., 2021) but was rapidly cleared. Emerging evidence suggests higher human-equivalent doses (120–180 mg/kg) may enhance efficacy (Campos et al., 2013, 2019; Li et al., 2023) emphasizing dose optimization for plausible prenatal care. Safety profiles and postnatal outcomes require further assessment.

In summary, our current study provides evidence linking genetic variation at *SERPINA1* Pi\**Z* with variation in gestational duration and the risk of SPTB. Our data supports a dual role for AAT in pregnancy: reducing inflammation-induced SPTB in deficient individuals and potentially protecting fetal growth and development. Our results suggest that exogenous AAT may cross the maternal-fetal interface (see Figure 8), although its exact protective mechanism against imminent inflammation-induced PTB remains elusive. It is likely that AAT’s effects may involve preservation of placental integrity, including stabilization of the extracellular matrix and support of trophoblast function, as suggested previously (Tiensuu et al., 2022). The therapeutic window appears narrow, with risks posed by both deficiency and excess AAT. AAT’s benefits may extend beyond acute cytokine modulation to broader immunological and placental pathways, as also suggested by epidemiological study of Orimoloye et al. (2024).

Future research should clarify tissue-specific AAT regulatory networks, optimize supplementation strategies, evaluate maternal–fetal transfer of exogenous AAT, assess long-term offspring outcomes, and explore applications in other pregnancy complications, including preeclampsia (Cosio et al., 2016; Feng et al., 2016). Present data and emerging new evidence underscore the importance of understanding systemic AAT deficiency as a risk factor for adverse pregnancy outcomes, with potential relevance to placental and fetal development.

## Conclusions

In present study we discovered that genetic AAT deficiency is a strong risk factor of SPTB. Very premature delivery is associated with chronic neurodevelopmental and pulmonary diseases. Our transgenic mice study lays a strong foundation for translational research harnessing AAT as a therapy for inflammation-mediated pregnancy disorders. Given the clinical approval of AAT augmentation therapy for specific lung and liver diseases, exploring its potential prenatal and perinatal applications holds significant promise.

## Data Availability

All data produced in the present work are contained in the manuscript.

## Acknowledgements

We would like to acknowledge Grifols for their generous sponsorship of this project through their research-sponsored program.

We want to acknowledge the participants and investigators of the FinnGen study. The FinnGen project is funded by two grants from Business Finland (HUS 4685/31/2016 and UH 4386/31/2016) and the following industry partners: AbbVie Inc., AstraZeneca UK Ltd, Biogen MA Inc., Bristol Myers Squibb Inc. (and Celgene Corporation & Celgene International II Sàrl), Genentech Inc., Merck Sharp & Dohme LCC, Pfizer Inc., GlaxoSmithKline Intellectual Property Development Ltd., Sanofi US Services Inc., Maze Therapeutics Inc., Johnson&Johnson Innovative Medicine Inc., Novartis AG, Boehringer Ingelheim International GmbH and Bayer AG. Following biobanks are acknowledged for delivering biobank samples to FinnGen: Auria Biobank (www.auria.fi/biopankki), THL Biobank (www.thl.fi/biobank), Helsinki Biobank (www.helsinginbiopankki.fi), Biobank Borealis of Northern Finland (https://www.ppshp.fi/Tutkimus-ja-opetus/Biopankki/Pages/Biobank-Borealis-briefly-in-English.aspx), Finnish Clinical Biobank Tampere (www.tays.fi/en-US/Research_and_development/Finnish_Clinical_Biobank_Tampere), Biobank of Eastern Finland (www.ita-suomenbiopankki.fi/en), Central Finland Biobank (www.ksshp.fi/fi-FI/Potilaalle/Biopankki), Finnish Red Cross Blood Service Biobank (www.veripalvelu.fi/verenluovutus/biopankkitoiminta), Terveystalo Biobank (www.terveystalo.com/fi/Yritystietoa/Terveystalo-Biopankki/Biopankki/) and Arctic Biobank (https://www.oulu.fi/en/university/faculties-and-units/faculty-medicine/northern-finland-birth-cohorts-and-arctic-biobank). All Finnish Biobanks are members of BBMRI.fi infrastructure (https://www.bbmri-eric.eu/national-nodes/finland/). Finnish Biobank Cooperative-FINBB (https://finbb.fi/) is the coordinator of BBMRI-ERIC operations in Finland. The Finnish biobank data can be accessed through the Fingenious^®^ services (https://site.fingenious.fi/en/) managed by FINBB.

We thank Maarit Haarala (University of Oulu, Oulu, Finland) for technical assistance.

We gratefully acknowledge the staff of the Oulu Laboratory Animal Centre (OULAC, Oulu, Finland) for their excellent care of the animals and their support throughout the *in vivo* experiments, as well as Hanna-Marja Voipio and Sakari Laaksonen for their advisory role during the license application. In this context, we would also like to acknowledge the Biocenter OULU (BCO) Transgenic and Tissue Phenotyping Core Facility (BCO-TTP) for their assistance in establishing our transgenic mouse lineage.

Furthermore, CSC–IT Center for Science, Finland, is acknowledged for computational resources.

## Data availability

All data produced in the present study are available upon reasonable request to the authors.

