## Supplementary figures for "Association of AAT/*SERPINA1* PI*Z Variant with Gestational Duration and Prevention of Premature Birth in Knockout Mice by AAT-Supplementation"

### Supplementary figures (in order of mention)

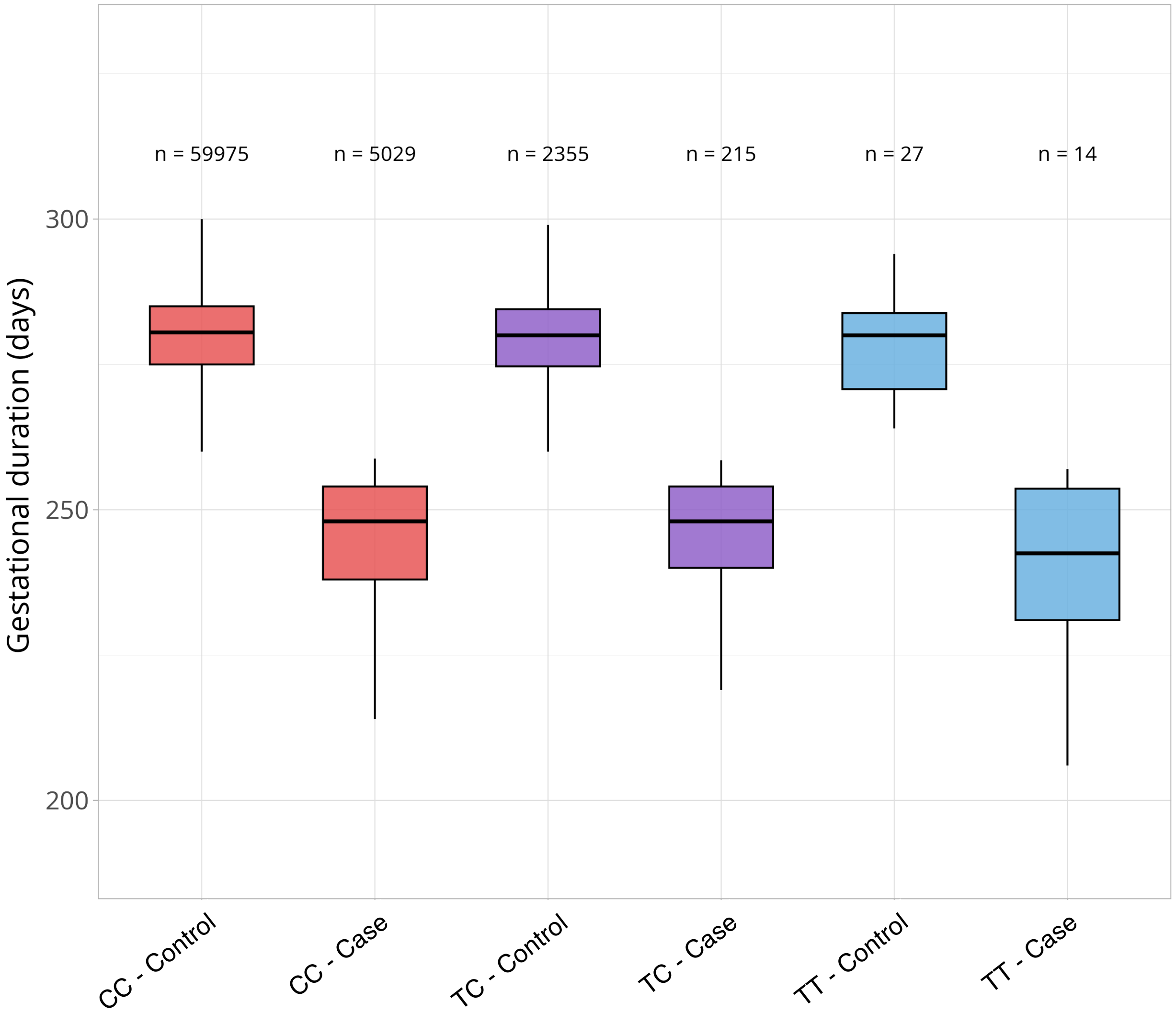


**Supplementary Figure 1. *SERPINA1* Pi*Z variant (rs28929474) genotypes (CC, TC and TT) and gestational duration in cases and controls.**

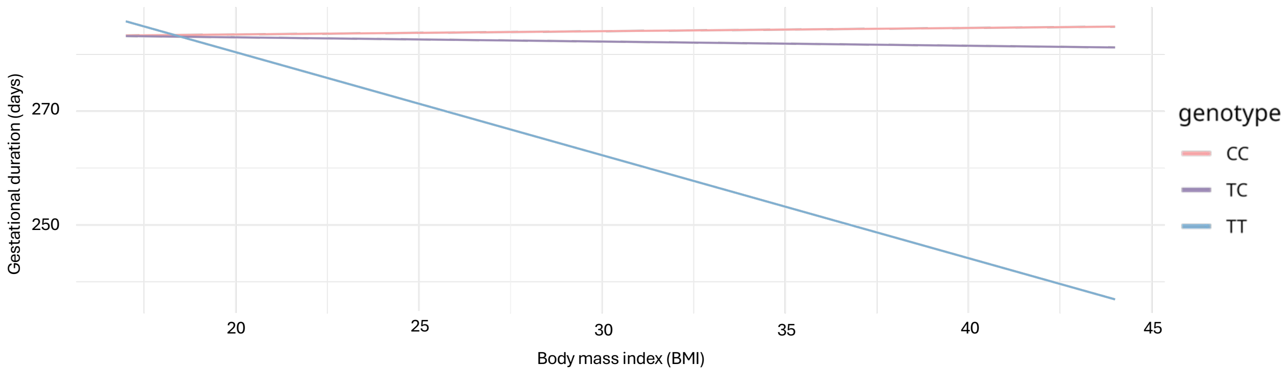


**Supplementary Figure 2. Gene x BMI interaction plot.** Increasing BMI (body mass index) was associated with shorter gestational duration among people with Serpina1 PIZZ genotype.

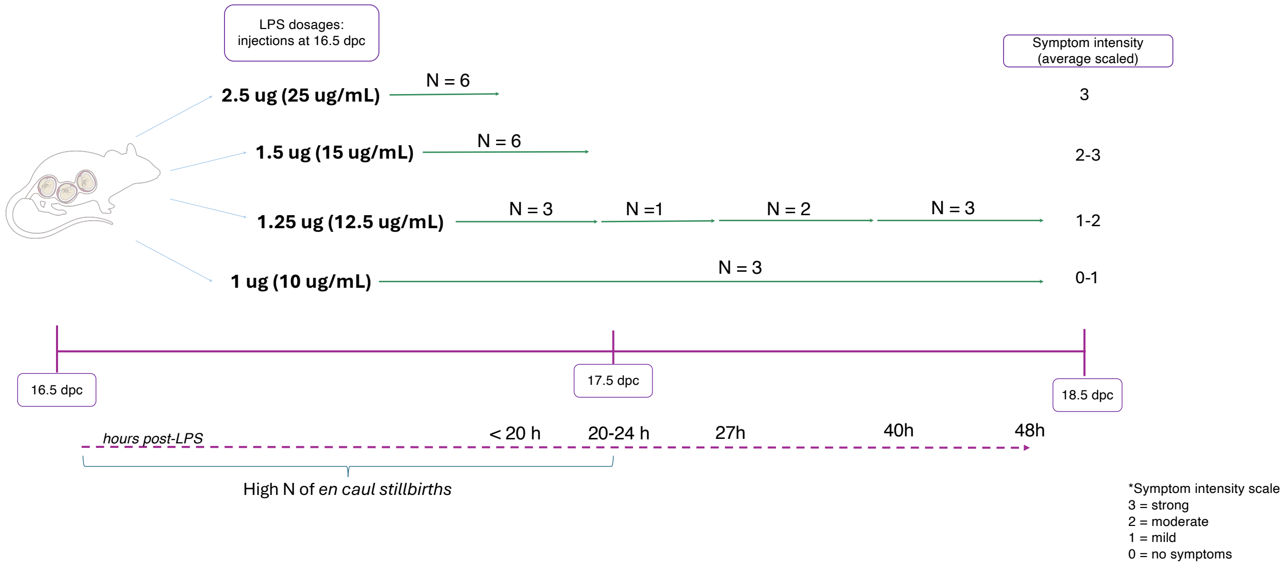


**Supplementary figure 3.** ***Serpina1a–e* knock-out and wild-type LPS dose testing**. Lipopolysaccharide (LPS) dosage was tested using four different doses: 1 µg (10 µg/mL), 1.25 µg (12.5 µg/mL), 1.5 µg (15 µg/mL), and 2.5 µg (25 µg/mL). Symptoms observed after administration included panting, disorientation, lethargy, eye squinting, and agitation. Symptom intensity was scored on a scale from 0–3 (0 = no symptoms; 1 = mild symptoms; 2 = moderate symptoms; 3 = strong symptoms). The number of births occurring by the indicated time point (either hours post-LPS or dpc) is shown as (N). Overall, the 2.5 µg (total N = 6) and 1.5 µg (total N = 6) doses resulted in the highest rates of preterm birth (PTB) and en caul stillbirths, suggesting strong inflammatory potency. The 1.25 µg dose (total N = 9) showed the most variable birth outcomes and was therefore selected as the most suitable dose for further experiments. The 1 µg dose (total N = 3) was considered too mild, as all dams tolerated it well and did not deliver preterm; thus, the number of animals was kept low to minimize animal use. Additional animals were included in the 1.25 µg group due to repeated testing performed to confirm its effect.

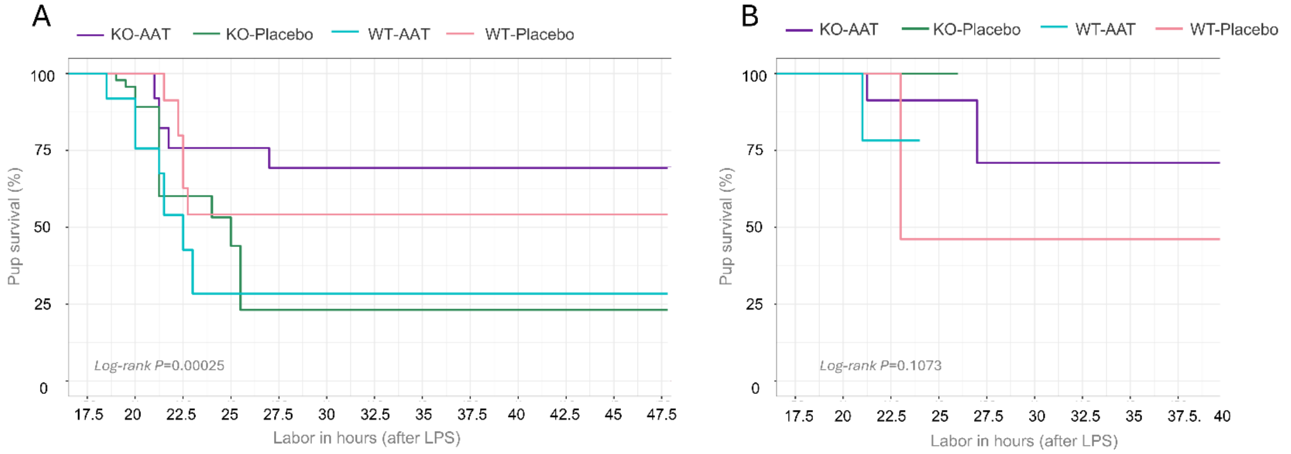


**Supplementary figure 4. Separate global Kaplan–Meier survival analysis of in utero (A) and ex utero (B) pups**. Dams in each group were treated with either placebo (KO-Placebo: green; WT-Placebo: pink) or Prolastin® (KO-AAT: purple; WT-AAT: turquoise), followed by lipopolysaccharide (LPS) administration. In utero viability was defined as pups remaining viable in the uterus at the time of dam euthanasia following the birth of the first pup. Ex utero viability was defined as pup survival after vaginal birth (excluding cesarean sections). The x-axis represents time in hours after LPS injection; each drop in the survival curve corresponds to the birth of a pup (end of observation for that individual). Survival analysis revealed a statistically significant difference among treatment groups in in utero survival (A) (Log-rank adjusted P = 0.00025), with the highest survival probability (>70%) observed in pups from Prolastin®-treated KO dams.

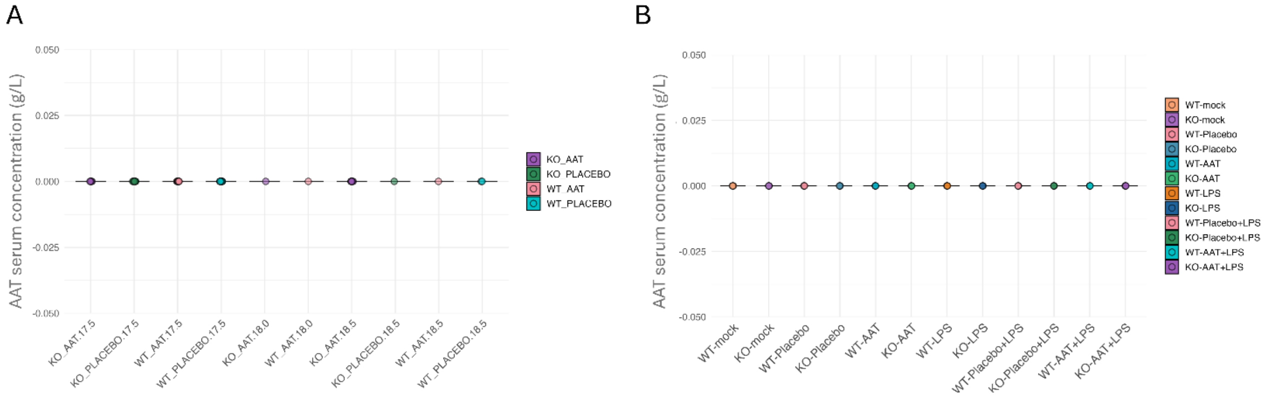


**Supplementary Figure 5. Human AAT serum concentration (g/L) from pups of Placebo and Prolastin®-treated dams.** Pups of Prolastin®/Placebo-tretated dams from cohort1 (A) or cohort2 (B) indicated no detectable serum levels of human derived AAT.

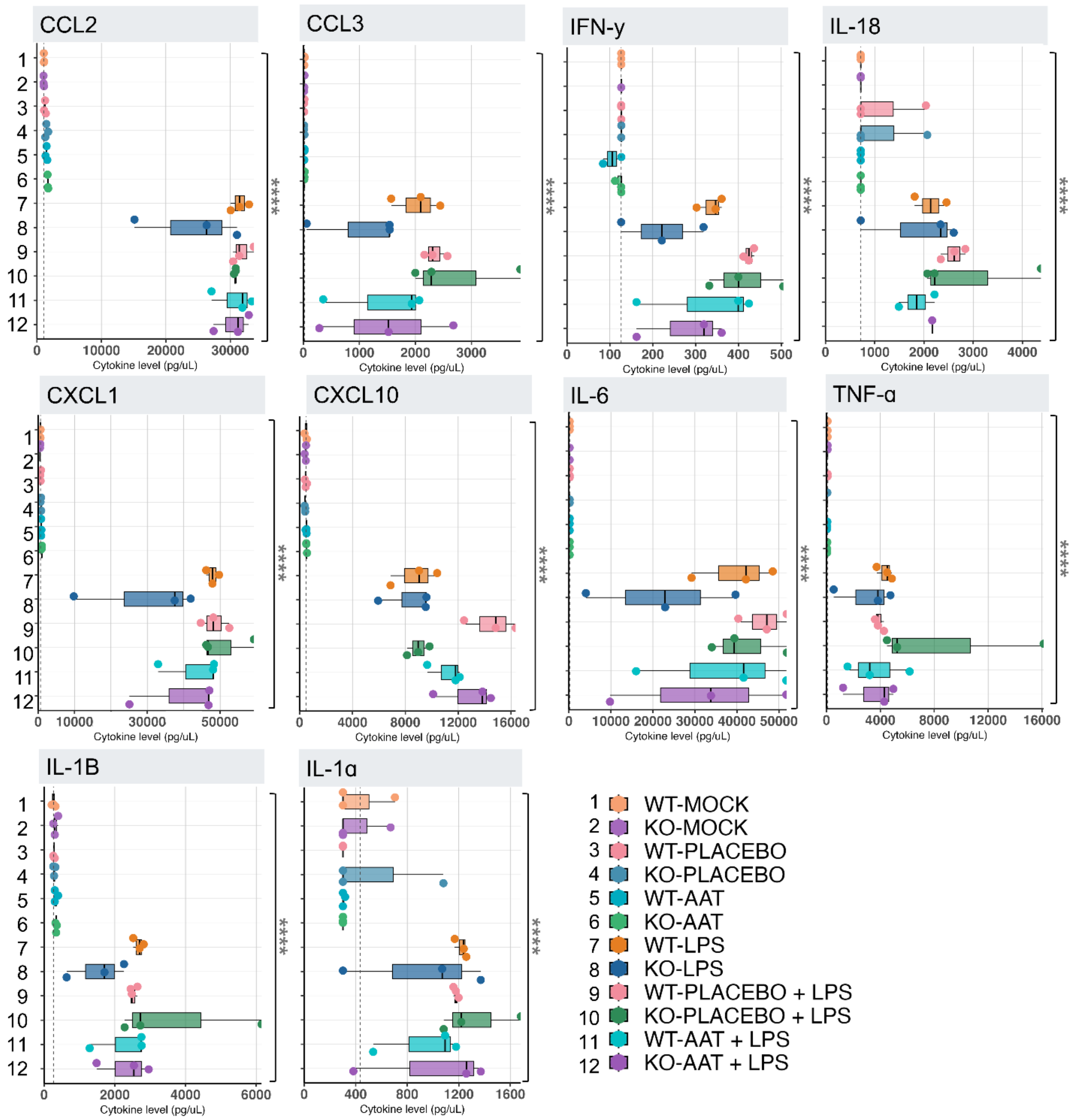


**Supplementary figure 6. Concentration (pg/uL) of pro-inflammatory-associated cytokines and factors.** C-C motif chemokine ligand 2 (CCL2), C-C motif chemokine ligand 3 (CCL3), C-X-C motif chemokine ligand 1 (CXCL1), C-X-C motif chemokine ligand 10 (CXCL10), interferon gamma (IFN-γ), interleukin-18 (IL-18), interleukin-1 beta (IL-1β), interleukin-1 alpha (IL-1α), interleukin-6 (IL-6), and tumor necrosis factor alpha (TNF-α), were quantified from maternal serum from *Placebo*- or Prolastin®- (AAT) treated dams with or w/o LPS, or LPS alone, at 16.5 dpc. Treatment groups (1. WT-mock; 2. KO-mock; 3. WT-*Placebo*; 4. KO-*Placebo*; 5. WT-AAT; 6. KO-AAT; 7. WT-LPS; 8. KO-LPS; 9. WT-*Placebo*+LPS; 10. KO-*Placebo*+LPS; 11. WT-AAT+LPS; 12. KO-AAT+LPS) plotted in boxplots. Wild-type mock group set as the visual reference (grey dashed line), with dots representing biological replicates (n=3/group). Error bars indicate the standard error (SE). LPS robustly induced a broad pro-inflammatory signature in both WT and KO mice, with significantly heightened responses (****, $P < 0.0001$) compared to non-LPS treated animals. These cytokines and chemokines are critical mediators in parturition-associated immune activation and are implicated in preterm labor pathogenesis. Prolastin®- treatment did not significantly reduce their expression compared to their Placebo-treated counterparts Statistical significance was assessed using individual Wilcoxon rank-sum tests (Mann–Whitney U tests) for binary treatment comparisons (e.g., LPS vs. no LPS) for each cytokine. Benjamini–Hochberg (BH) correction was applied to adjust for multiple testing.

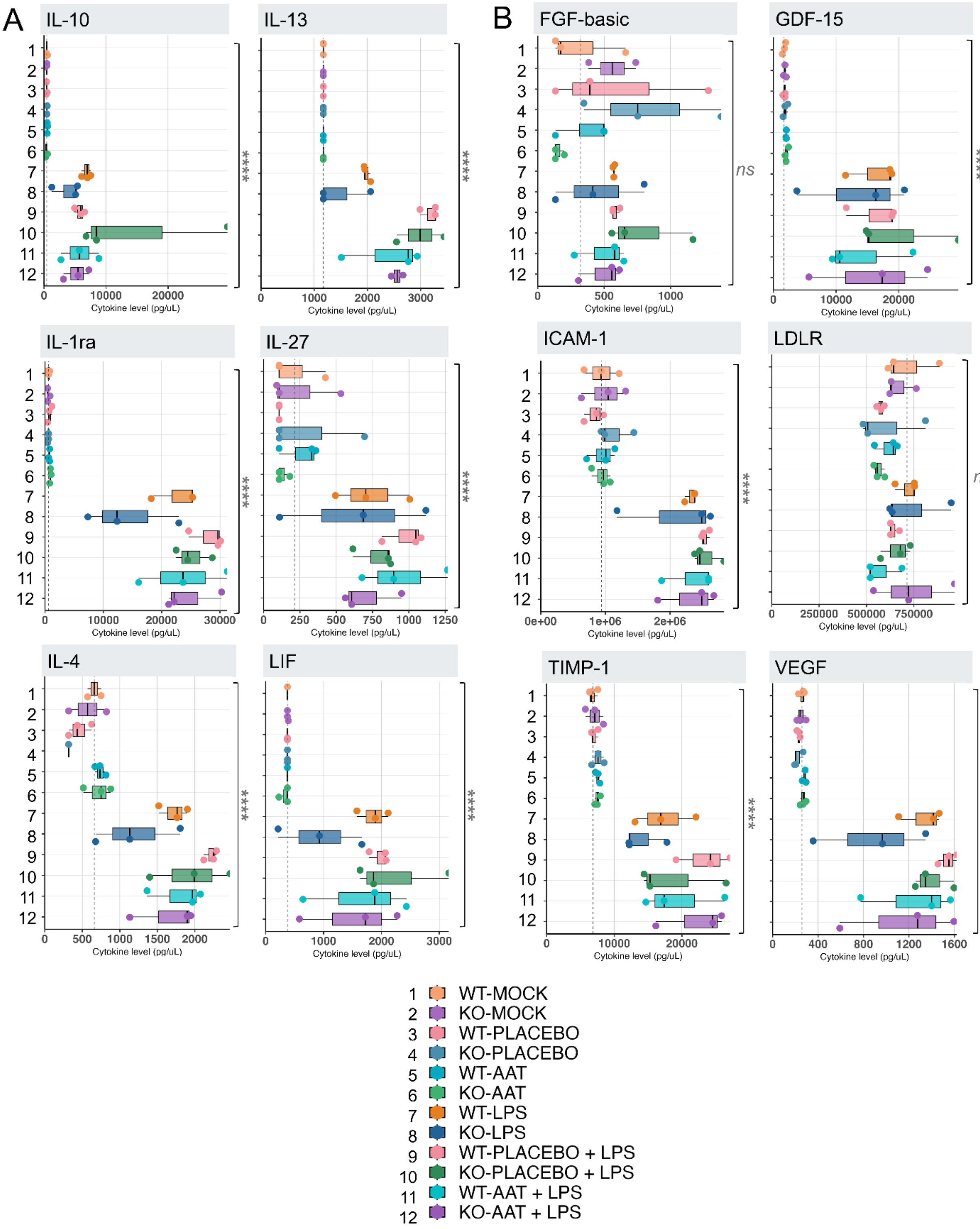


**Supplementary figure 7**. **Concentration (pg/uL) of anti-inflammatory cytokines**. Interleukin-10 (IL-10), interleukin-1 receptor antagonist (IL-1ra), interleukin-4 (IL-4), interleukin-13 (IL-13), leukemia inhibitory factor (LIF), and interleukin-27 (IL-27) **(A)**, and growth and proliferation-associated cytokines and factors, fibroblast growth factor-basic (FGF-basic), growth differentiation factor 15 (GDF-15), intercellular adhesion molecule-1 (ICAM-1), low-density lipoprotein receptor (LDLR), tissue inhibitor of metalloproteinases-1 (TIMP-1), and vascular endothelial growth factor (VEGF) **(B)**, were quantified from maternal serum from *Placebo*- or Prolastin®- (AAT) treated dams with or w/o LPS, or LPS alone, at 16.5 dpc. Treatment groups (1. WT-mock; 2. KO-mock; 3. WT-*Placebo*; 4. KO-*Placebo*; 5. WT-AAT; 6. KO-AAT; 7. WT-LPS; 8. KO-

LPS; 9. WT-*Placebo*+LPS; 10. KO-*Placebo*+LPS; 11. WT-AAT+LPS; 12. KO-AAT+LPS) plotted in boxplots. Wild-type mock group set as the visual reference (grey dashed line), with dots representing biological replicates (n=3/group). Error bars indicate the standard error (SE). While LPS stimulated expression of several anti-inflammatory mediators, the magnitude and pattern of induction varied by genotype. LPS injection significantly elevated the expression of all the presented anti-inflammatory mediators (****, P <0.0001). In addition, LPS exposure significantly increased VEGF, TIMP-1, ICAM-1, and GDF-15 levels (P < 0.0001). AAT treatment did not however significantly affect none of the anti-inflammatory or remodeling-associated factors. Statistical significance was assessed using individual Wilcoxon rank-sum tests (Mann–Whitney U tests) for binary treatment comparisons (e.g., LPS vs. no LPS) for each cytokine. Benjamini–Hochberg (BH) correction was applied to adjust for multiple testing.

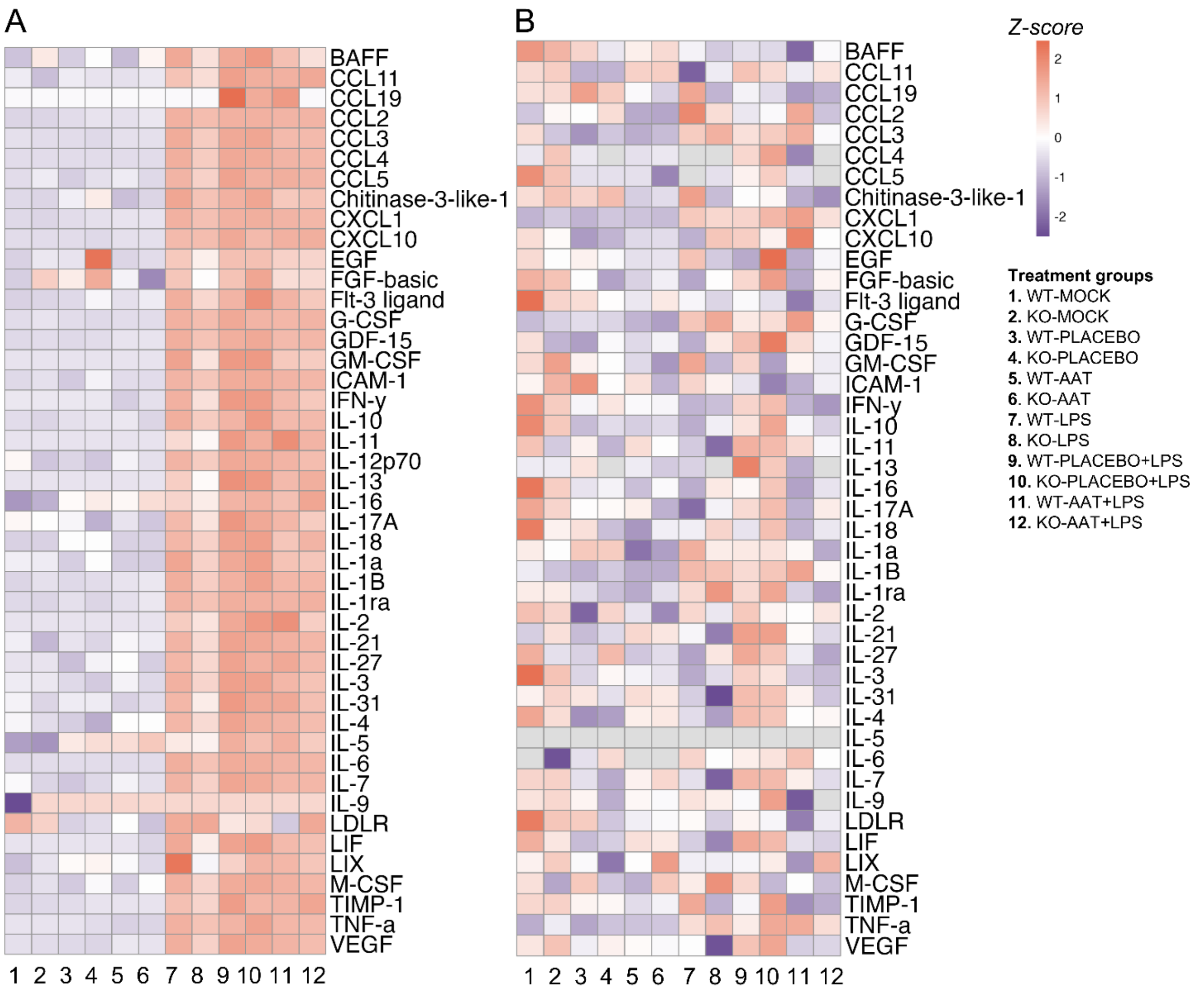


**Supplementary figure 8. Z-score heatmaps showing cytokine and chemokine profiles in maternal (A) and fetal (B) serum at 16.5 dpc following placebo or AAT treatment with or without LPS exposure**. Maternal profiles showed clear separation between LPS (7-12) and non-LPS groups (1-6), while fetal serum profiles for these analytes exhibited no distinct pattern based on dam genotype or treatment.

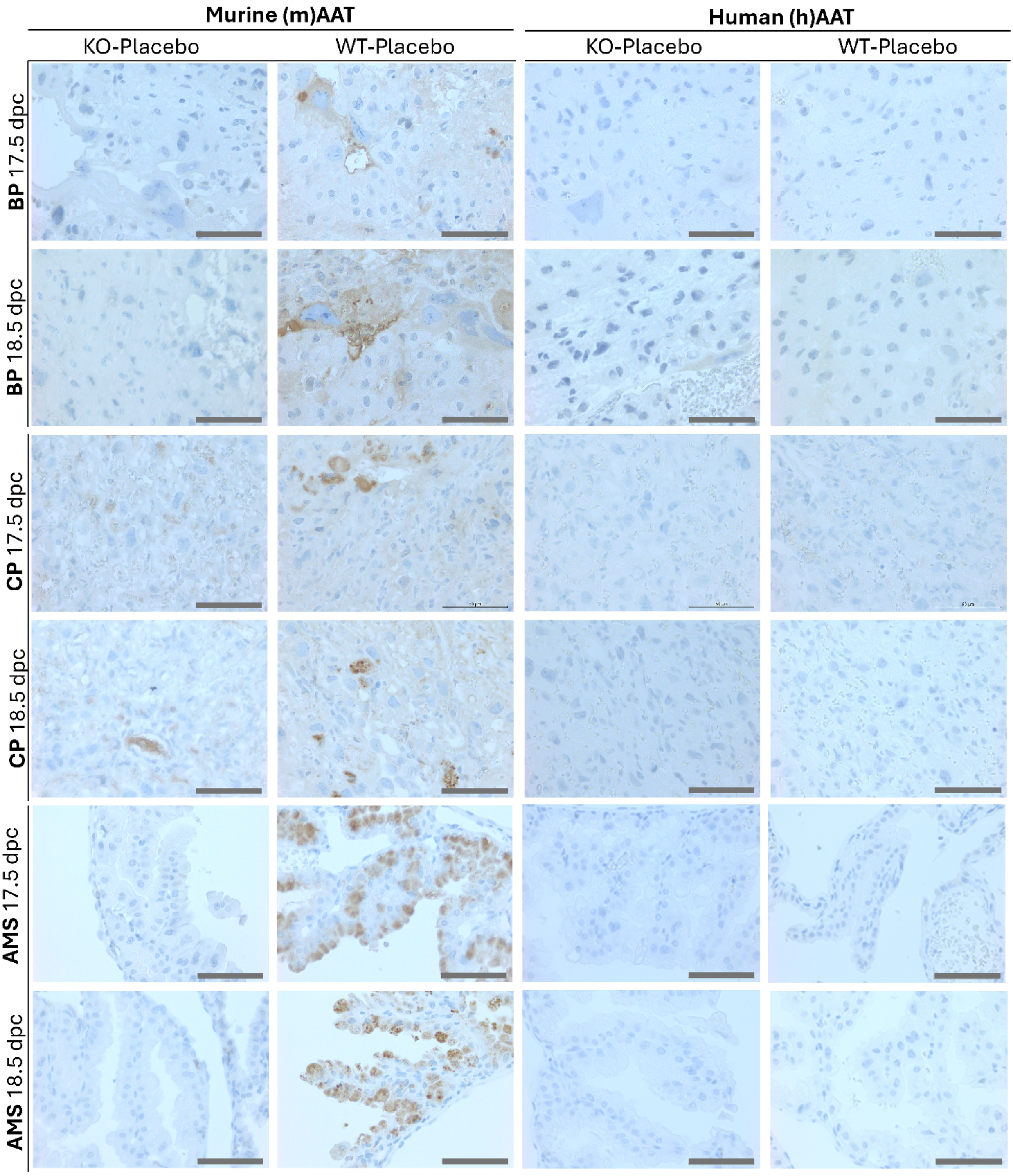


**Supplementary figure 9. Immunohistochemistry (IHC) of placental samples from placebo-treated *Serpina1a–e* knockout (KO) dams at 17.5 and 18.5 days post coitum (dpc).** Representative IHC images of placental sections from KO and WT dams at 17.5 dpc (preterm) and 18.5 dpc (term) are shown. Anatomical structures including the basal plate (BP), chorionic plate (CP), and amniotic membranes (AMS) are labeled in the left panels along with the corresponding gestational day. Staining with mouse AAT (mAAT; two leftmost columns) and human AAT (hAAT; two rightmost columns) antibodies is indicated at the top of the panels. In KO-Placebo samples, mAAT staining was absent in the BP and limited in the AMS. The CP exhibited localized staining, likely reflecting endogenous AAT expression from heterozygous pups. In contrast, WT-Placebo placentas showed more widespread and consistent mAAT expression across all regions and timepoints. As expected, hAAT staining was absent in both KO and WT placebo-treated placental sections, confirming the lack of exogenous human AAT in these samples.

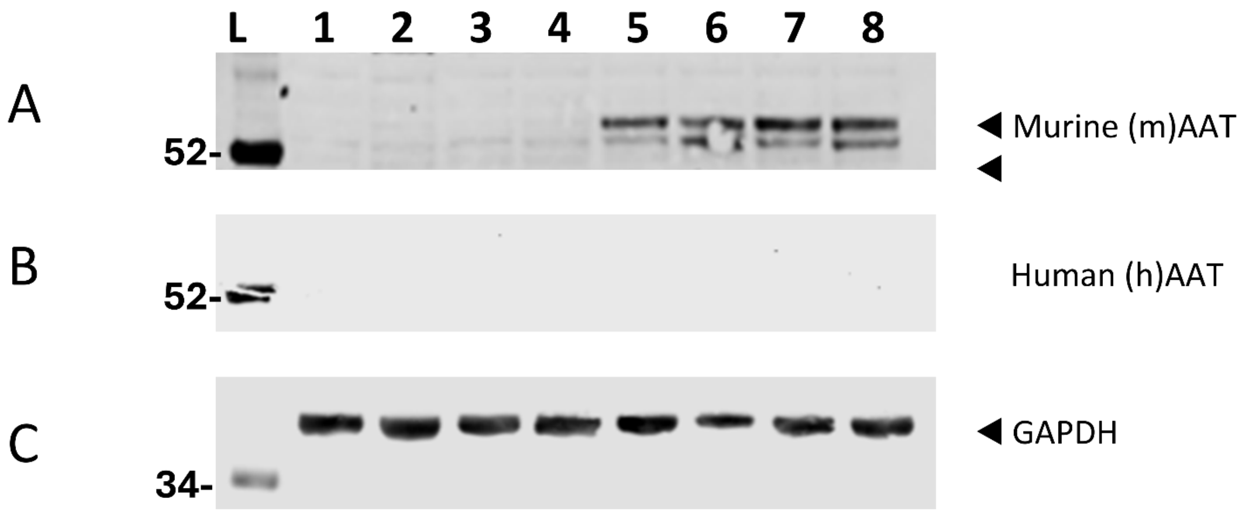


**Supplementary figure 10. Western blot analysis of liver samples from 17.5 and 18.5 days post coitum (dpc) Serpina1a–e knockout (KO) and wild-type (WT) dams.** Western blot (WB) analysis was performed on liver samples from Prolastin® (human alpha-1 antitrypsin, hAAT)- and placebo-treated dams at 17.5 dpc (preterm birth: PTB) and 18.5 dpc (term birth: TB). Band formation for mouse AAT (mAAT) is shown in panel (A), hAAT in panel (B), and the housekeeping protein glyceraldehyde 3-phosphate dehydrogenase (GAPDH) in panel (C). No hAAT bands were detected in any of the samples. Sample order in lanes 1–8 (including molecular weight ladder [L]) is as follows:1) KO-Placebo, PTB; 2. KO-Placebo, TB; 3. KO-AAT, PTB; 4. KO-AAT, TB; 5. WT-Placebo, PTB; 6. WT-Placebo, TB; 7. WT-AAT, PTB; 8. WT-AAT, TB. Molecular weight markers in the ladder are indicated at 52 kilodaltons (kDa) and 34 kDa. The mAAT bands at approximately 52 kDa and ~54 kDa likely represent two glycosylated isoforms of AAT (partially and fully glycosylated). GAPDH band formation at ~37 kDa is within the expected range.

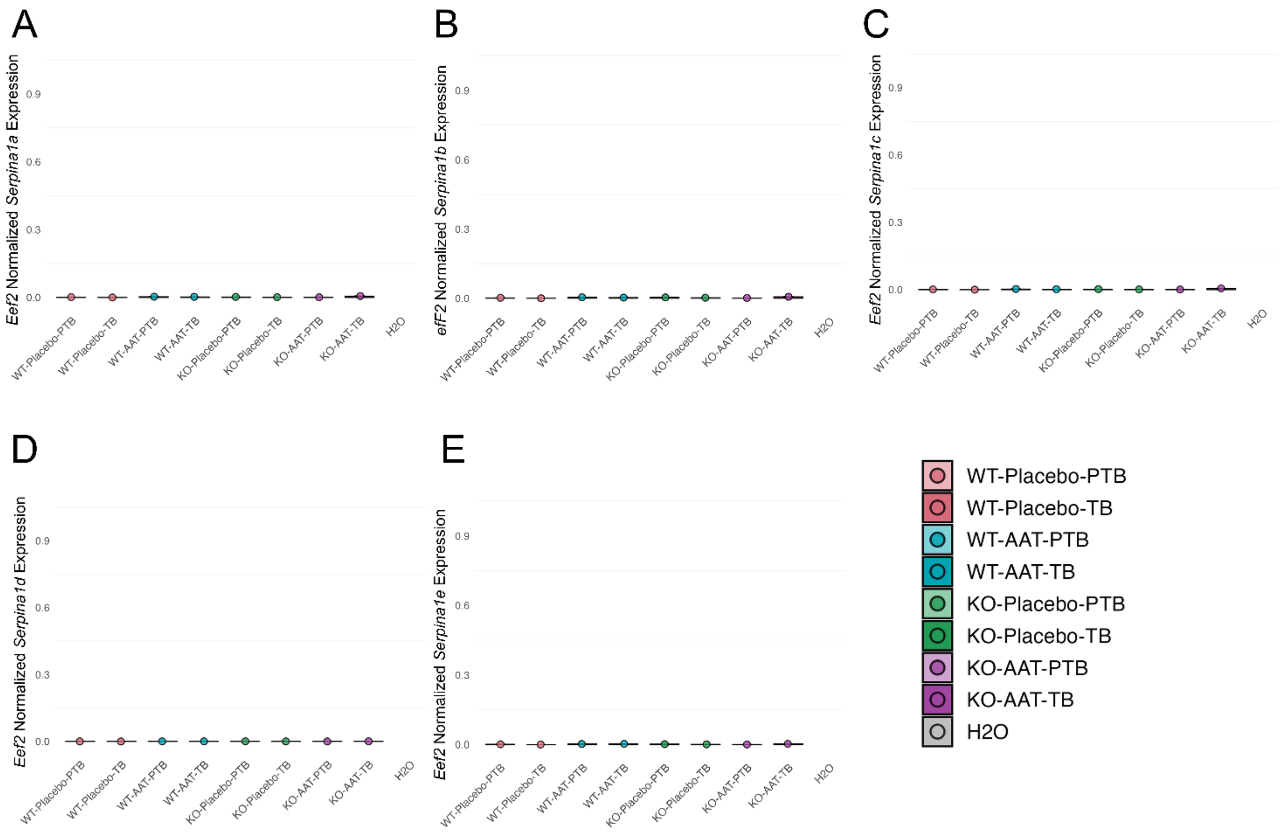


**Supplementary Figure 11. Placental mRNA expression of *Serpina1a–e* paralogs at 17.5 dpc (PTB) and 18.5 dpc (TB)** from cohort 1 (same animals as in Figure 3). Plots (A–E) show individual *Serpina1* paralog (*a–e*) expression by treatment group, normalized to *Eef2*. No detectable transcripts were quantified for any of the paralogs in any of the samples.
